# The link between the *ANPEP* gene and type 2 diabetes mellitus may be mediated by the disruption of glutathione metabolism and redox homeostasis

**DOI:** 10.1101/2024.03.16.24304385

**Authors:** Yaroslava Korvyakova, Iuliia Azarova, Elena Klyosova, Maria Postnikova, Victor Makarenko, Olga Bushueva, Maria Solodilova, Alexey Polonikov

## Abstract

Aminopeptidase N (ANPEP), a membrane-associated ectoenzyme, has been identified as a susceptibility gene for type 2 diabetes (T2D) by genome-wide association and transcriptome studies; however, the mechanisms by which this gene contributes to disease pathogenesis remain unclear. The aim of this study was to determine the comprehensive contribution of ANPEP polymorphisms to T2D risk and annotate the underlying mechanisms. A total of 3206 unrelated individuals including 1579 T2D patients and 1627 controls were recruited for the study. Twenty-three common functional single nucleotide polymorphisms (SNP) of ANPEP were genotyped by the MassArray-4 system. Six polymorphisms, rs11073891, rs12898828, rs12148357, rs9920421, rs7111, and rs25653, were found to be associated with type 2 diabetes for the first time (P_perm_≤0.05). Common haplotype rs9920421G-rs4932143G-rs7111T was strongly associated with increased risk of T2D (P_perm_=5.9×10^-12^), whereas two rare haplotypes such as rs9920421G-rs4932143C-rs7111T (P_perm_=6.5×10^-40^) and rs12442778A-rs12898828A-rs6496608T-rs11073891C (P_perm_=1.0×10^-7^) possessed strong protection against disease. We identified 38 and 109 diplotypes associated with T2D risk in males and females, respectively (FDR≤0.05). ANPEP polymorphisms showed associations with plasma levels of fasting blood glucose, aspartate aminotransferase, total protein and glutathione (P<0.05), and several haplotypes were strongly associated with the levels of reactive oxygen species and uric acid (P<0.0001). A deep literature analysis has facilitated the formulation of a hypothesis proposing that increased plasma levels of ANPEP as well as liver enzymes such as aspartate aminotransferase, alanine aminotransferase and gamma-glutamyltransferase serve as an adaptive response directed towards the restoration of glutathione deficiency in diabetics by stimulating the production of amino acid precursors for glutathione biosynthesis.

**Highlights:**

- Polymorphisms, rs11073891, rs12898828, rs12148357, rs9920421, rs7111, and rs25653 of *ANPEP*, were discovered as novel susceptibility markers for type 2 diabetes mellitus.
- ne common and two rare haplotypes of *ANPEP* are strongly associated with increased and decreased risk of type 2 diabetes, respectively.
- Thirty eight and one hundred and nine *ANPEP* diplotypes influenced disease risk in males and females, respectively.
- Polymorphisms of *ANPEP* were associated with plasma levels of fasting blood glucose, aspartate aminotransferase, total protein, glutathione, reactive oxygen species and uric acid in diabetics.
- Increased plasma levels of ANPEP and liver enzymes (AST, ALT, and GGT) in T2D patients may reflect an adaptive response to glutathione deficiency through the formation of more amino acid precursors of glutathione.

## 1. Introduction

The International Diabetes Federation reported in 2021 that approximately 537 million adults aged 20–79 years, or 10.5% of the global population, were estimated to have diabetes [1]. Type 2 diabetes (T2D) mellitus accounts for more than 90% of all diabetes cases and is attributed to defective insulin secretion by pancreatic β-cells and the inability of insulin-sensitive tissues to respond appropriately to insulin [2,3]. Pancreatic β-cell is the primary regulatory center controlling glucose homeostasis and the impairment of β-cells caused by islet reduced number and/or reduced number of cells is thought to be a central disorder responsible for the development and progression of type 2 diabetes [3,4,5]. The majority of T2D cases have a multifactorial origin, meaning that multiple genetic, behavioral and environmental risk factors jointly contribute to disease development and progression [6,7,8,9]. Environmental risk factors for T2D include consumption of energy-dense food, sedentary lifestyle, and aging [10,11], and numerous single nucleotide polymorphisms (SNP) have been found to be associated with disease susceptibility [7,12,13]. Frequently, impairment of insulin secretion and defects in insulin action coexist in the same patient, making it uncertain which abnormality, if either alone, is the primary cause of hyperglycemia [14]. Although endoplasmic reticulum stress, lipotoxicity, glucotoxicity, mitochondrial dysfunction, islet inflammation and oxidative stress, have been established as factors that contribute to decreased pancreatic β-cell function in T2D [15–20], the primary factors and molecular mechanisms initiating progressive β-cell failure remain unknown.

We recently proposed that a primary pathological condition responsible for oxidative stress and induces cellular stress responses, leading to pancreatic β-cell death and type 2 diabetes mellitus, is an endogenous deficiency of glutathione [21–23]. Glutathione, a tripeptide consisting of three amino acids: glutamate, cysteine, and glycine, is the most abundant non-protein thiol in cells, acting as the principal reducing agent and potent antioxidant, protecting the cell from oxidative damage [24]. Reduced glutathione (GSH) levels within cells are maintained at their highest (millimolar) level, suggesting their importance beyond antioxidant defense [24–26]. It also executes an array of biological functions, including xenobiotic and endogenous toxic substance detoxification, mitochondrial redox balance maintenance, antiviral defense, immune response, regeneration of vitamin C and E, regulation of cell proliferation, apoptosis, and protein folding [24–26]. The hypothesis that endogenous glutathione deficiency is the primary cause of type 2 diabetes is supported by several research findings. First, glutathione deficiency, both in the blood plasma and within cells in patients with T2D, is regularly reported in the literature [27–34]. Second, many known environmental risk factors for type 2 diabetes were found to be responsible for glutathione deficiency through decreased synthesis and increased utilization of GSH, or a combination of both [22]. Third, decreased glutathione levels are closely correlated with the pathological conditions found in T2D, and replenishment of glutathione deficiency improves them. These pathological changes include oxidative stress and mitochondrial dysfunction [31,32,35–37], β-cell dedifferentiation and failure [38], impaired glucose tolerance, insulin secretion [39–41] and sensitivity [42,43]. In addition, the depletion of endogenous glutathione is correlated with increased blood glucose levels and contributes to the complications of type 2 diabetes [33,34,38]. Finally, additional evidence for glutathione deficiency in T2D is the results of genetic studies demonstrating a significant role of loss-of-function variants of genes encoding enzymes of glutathione metabolism in determining disease susceptibility [21,22,44–46], and the link of these genes to T2D risk is explained by decreased activity and/or expression of enzymes involved in glutathione biosynthesis, including in pancreatic beta cells [23,47].

ANPEP, an alanyl aminopeptidase, is a membrane-associated enzyme involved in glutathione metabolism with broad substrate specificity [48]. It catalyzes the hydrolysis of peptide bonds with the removal of amino acids from the amino termini of proteins and peptides. As a part of the glutathione metabolism pathway, ANPEP hydrolyzes the peptide L-cysteinylglycine to cysteine and glycine substrates for *de novo* biosynthesis of GSH [48,49]. A transcriptomic study performed on pancreatic beta cells has revealed *ANPEP* to be a differentially expressed (up-regulated) gene in patients with type 2 diabetes [50]. Moreover, a genome-wide association study in a South Asian population identified several SNPs located at chromosome 15 near the *ANPEP* and *AP3S2* genes that were associated with susceptibility to T2D [51]. Further study of Locke using allelic expression profiling showed that the T2D-related polymorphisms (for example, SNP rs2007084) are associated with increased expression of *ANPEP* in the pancreatic islets of diabetics [52]. Pedersen et al. [53] performed system biology and bioinformatics analyses by integrating islet gene and protein expression data with protein interactions in pancreatic islet protein complexes and identified *ANPEP* as one of the top overlapping genes closely associated with T2D susceptibility. Taken together, the above findings clearly show a plausible role for *ANPEP* in the pathogenesis of T2D; however, the mechanisms by which this gene contributes to T2D susceptibility remain unclear.

We hypothesize that *ANPEP* gene polymorphisms are associated with the risk of type 2 diabetes through the key role of this enzyme in the metabolism of glutathione. To date, no studies have comprehensively analyzed the role of functionally significant *ANPEP* polymorphisms in the development of type 2 diabetes mellitus. This has become the basis for a detailed analysis of the relationship between a wide range of functionally significant polymorphisms of the *ANPEP* gene and the risk of type 2 diabetes mellitus. Moreover, we conducted a comprehensive analysis of literature and online resources on biochemistry and metabolism related to ANPEP, which allowed us to annotate the potential molecular mechanisms by which the enzyme is involved in the pathogenesis of type 2 diabetes mellitus.

## 2. Materials and methods

### 2.1. Study participants and diagnosis of type 2 diabetes

The Kursk State Medical University Regional Ethics Review Committee approved the study protocol and adhered to the ethical guidelines outlined in the Declaration of Helsinki. Prior to enrollment, written informed consent was obtained from each participant. A total of 3206 unrelated Russians participated in the study, consisting of 1579 patients with T2D and 1627 age- and sex-matched healthy individuals in the control group. Most of the participants were from the Central Russia, Kursk region. Participants were interviewed in person using a questionnaire from one of our previous studies [54]. Between November 2016 and October 2019, the Endocrinology Division of Kursk Emergency Hospital admitted patients diagnosed with type 2 diabetes according to World Health Organization criteria [55], which included an HbA1c level of 6.5%, random blood glucose level of 11.1 mmol/L, or fasting blood glucose (FBG) level of 7.0 mmol/L. The inclusion criteria were previously described [56], and written informed consent was obtained from all the study participants. The clinical, laboratory, and demographic characteristics of the research groups were reported in our most recent paper [57].

### 2.2 Genetic analysis

Venous blood samples were obtained from the cubital veins of study participants and placed in EDTA-coated tubes. The samples were subsequently frozen and stored at −20°C until further processing. Phenol/chloroform extraction and ethanol precipitation have been used to purify total DNA. A total of twenty three single nucleotide polymorphisms in the *ANPEP* gene (Supplementary Table A) were selected for the study based on their functional properties and linkage disequilibrium (HapMap data, European population). In particular, the selection of SNPs was conducted using the Candidate Gene SNP Selection tool (GenePipe), available online at the SNPinfo Web Server (https://snpinfo.niehs.nih.gov/snpinfo/selegene.html). The MALDI-TOF mass spectrometry iPLEX platform on the MassArray-4 system (Agena Bioscience, Inc., San Diego, CA, USA) was used to genotype twenty SNPs whereas the CFX-96 Thermocycler (Bio-Rad, USA) was used for real-time PCR genotyping of three remaining SNPs (rs16943590, rs753362, and rs7111). The primer sequences used for genotyping are available upon request. To ensure quality control, 5% of the DNA samples were genotyped twice by unaware researchers, and the concordance rate was greater than 99%.

### 2.3 Biochemical analysis

The plasma levels of glutathione (GSH and GSSG) were determined in study patients using a fluorometric test method and GSH/GSSG Ratio Detection test Kit II (Abcam, USA). The concentration of reactive oxygen species (ROS) in plasma was measured using a fluorometric assay and OxiSelectTM In Vitro ROS/RNS Assay Kit (Cell Biolabs, USA) on the Varioscan Flash microplate reader (Thermo Fisher Scientific, USA). The plasma concentrations of glycated hemoglobin (HbA1c), glucose, total cholesterol, high- and low-density lipoproteins, triacylglycerol, uric acid, alanine transaminase (AST), alanine aminotransferase (ALT) and total protein in blood plasma were measured using a semi-automated biochemical analyzer (Clima MC-15, Ral Tecnica Para el Laboratorio, S.A., Spain) and reagents manufactured by DIAKON-DS (Moscow, Russia).

### 2.4 Statistical and bioinformatics analysis

The statistical power for the genetic association study was calculated using the Michigan Power Calculator (http://csg.sph.umich.edu/abecasis/gas_power_calculator/). Assuming an 85-90% power and a 5% type I error (which equals 0.05), the association study of selected polymorphisms with T2D risk could reveal a genotype relative risk of 1.26-1.51 based on the sample sizes of 1579 T2D cases and 1627 healthy controls. Differences in the genotype and allele frequencies between T2D patients and healthy controls were compared using chi-square tests. The PLINK software 1.9 [58] was used to examine allele and genotype frequencies in the study groups and their relationship with T2D risk using logistic regression analysis. The calculated odds ratio (OR) and 95% confidence intervals (95% CI) for disease risk were adjusted for age, sex, and body mass index (BMI). Allelic, recessive, dominant, and log-additive genetic models were assessed for SNP-disease relationships. The false discovery rate (FDR) procedure was used (False Discovery Rate Online Calculator, https://www.sdmproject.com/utilities/?show=FDR) to adjust for multiple comparisons. The Haploview program version 4.2 [59] was used to visualize the structure of the *ANPEP* haplotypes and assess their association with T2D risk by calculating P-values through adaptive permutations. The STATISTICA v.13 software was used to estimate the associations between *ANPEP* genotype combinations (diplotypes) and T2D risk. The normality of the biochemical parameters was examined using the Kolmogorov-Smirnov test, and Student’s t-test was used to compare the mean with standard deviation between the groups. Non-normally distributed characteristics were expressed in median (Me) with interquartile range (IQR) and compared between the groups using the Kruskal-Wallis test. To validate the associations of the studied gene variants with the risk of T2D, replication analysis was carried out in eleven populations using summary statistics from the UK Biobank population (http://geneatlas.roslin.ed.ac.uk) and genome-wide association studies published at the T2D Knowledge portal (http://www.type2diabetesgenetics.org). Functional annotation of the studies SNPs was performed using bioinformatics online resources such as eQTLGen Consortium (https://www.eqtlgen.org/phase1.html) and HaploReg v4.2 (https://pubs.broadinstitute.org/mammals/haploreg/haploreg.php)

## 3. Results

### 3.1 Association of the ANPEP gene polymorphisms with the risk of T2D

The genotype frequencies of the twenty three SNPs in the *ANPEP* gene were in Hardy-Weinberg equilibrium in both the case and control groups (P>0.05). A summary of the associations between polymorphisms and the risk of T2D is presented in Table 1 (the associations between SNPs and T2D in sex-stratified groups are shown in Supplementary Table B). Allelic, additive, dominant, and recessive genetic models were evaluated, and P-values were calculated using permutations. The most significant permutation P-value (P_perm_) was considered to select the best genetic model for SNP-disease association. The genotype and allele frequencies of the *ANPEP* gene in the study groups are shown in Table 2 (the genotype and allele frequencies of *ANPEP* in sex-stratified groups are shown in Supplementary Table 3). As seen in Table 2, seven polymorphisms (rs25653, rs12898828, rs11073891, rs12148357, rs9920421, rs4932143, and rs7111) were significantly associations with the risk of T2D. Sex-stratified analysis (Supplementary Table C) revealed that only two SNPs (rs11073891 and rs4932143) were associated with T2D in both sexes, while rs25653 was associated with T2D risk exclusively in males (OR=0.41, 95% CI=0.52-0.96, Pperm=0.02, recessive model). Four SNPs such as rs12898828, rs6496608, rs9920421, and rs7111 were significantly associated with the risk of T2D in females. In addition, two polymorphisms of *ANPEP* were found to be associated with the age of disease onset. Specifically, individuals with the rs17240268-A/A genotype (Me=44.5 IQR 40; 49) manifested T2D 7.5 years earlier than those with G/G (Me=52 IQR 45; 58) or G/A (Me=50 IQR 45; 57) genotypes (P=0.023). Moreover, females with genotype rs72756574-G/G (Me=48.5 IQR 45; 52) manifested T2D 1.5-3 years earlier than females with rs72756574-G/T (Me=50 IQR 46; 56) or rs72756574-T/T (Me=53 IQR 46; 59) genotypes (P=0.009).

**Table 1.**
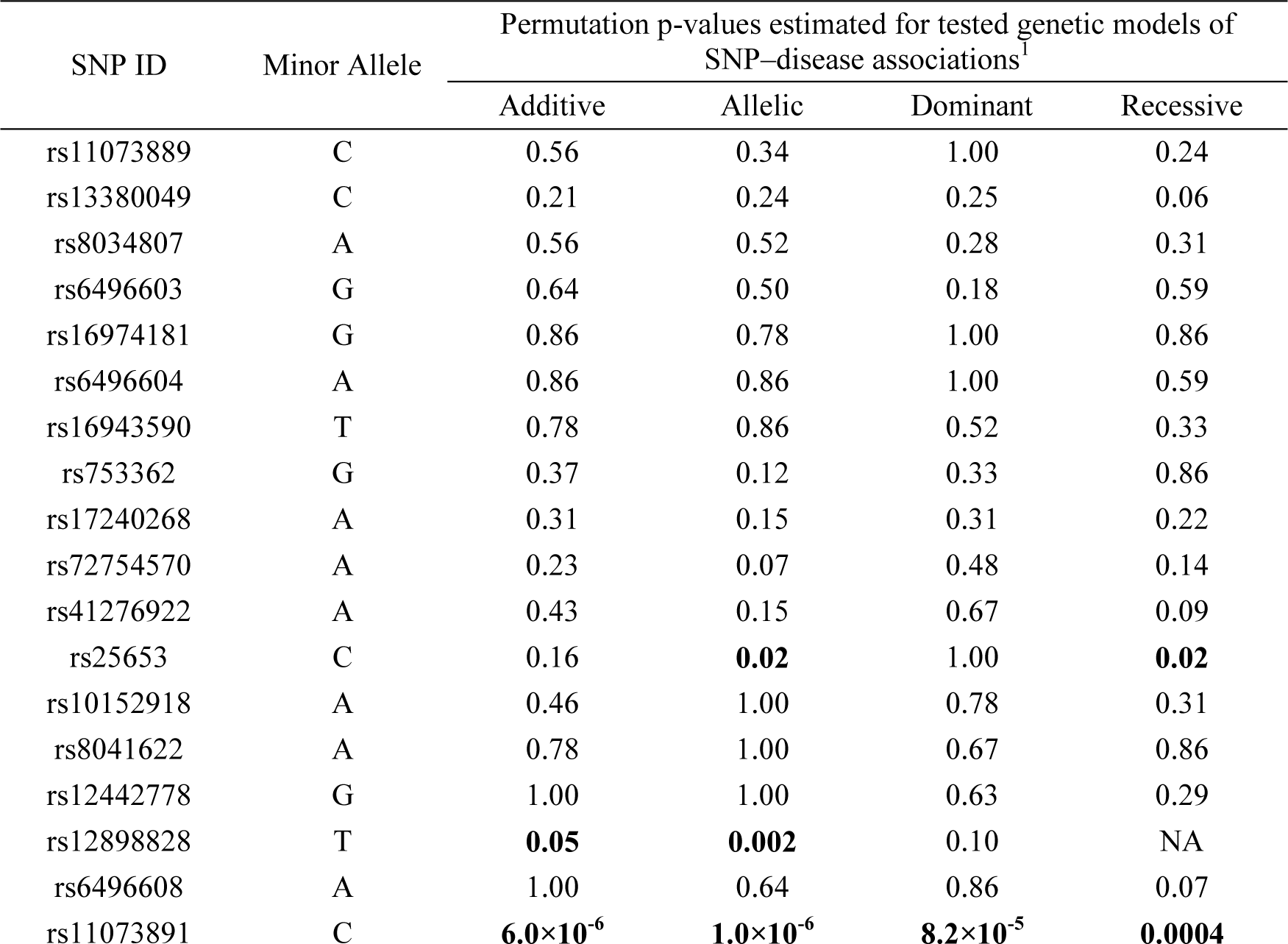
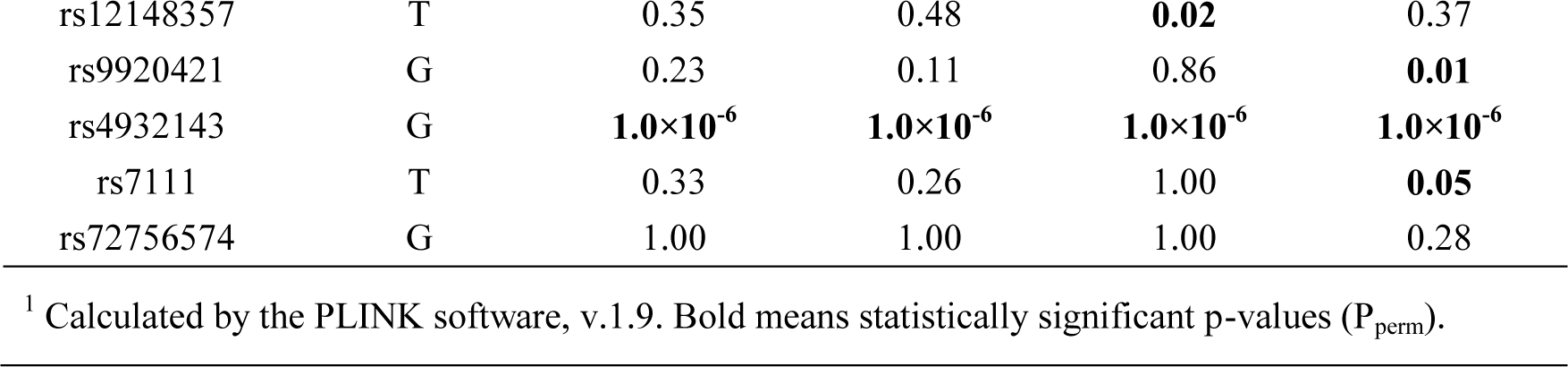
A summary of associations between *ANPEP* gene polymorphisms and T2D risk.

**Table 2.**
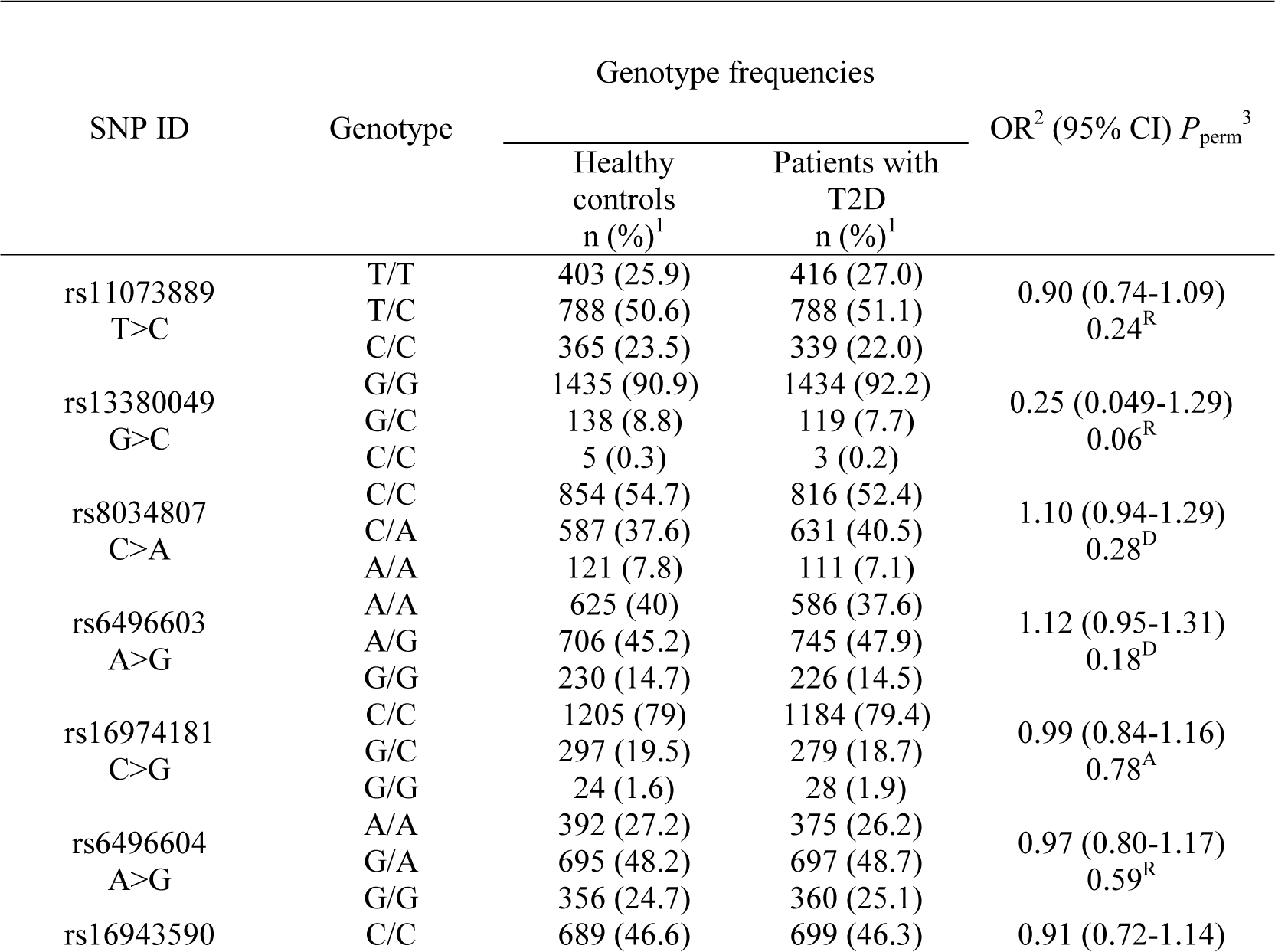
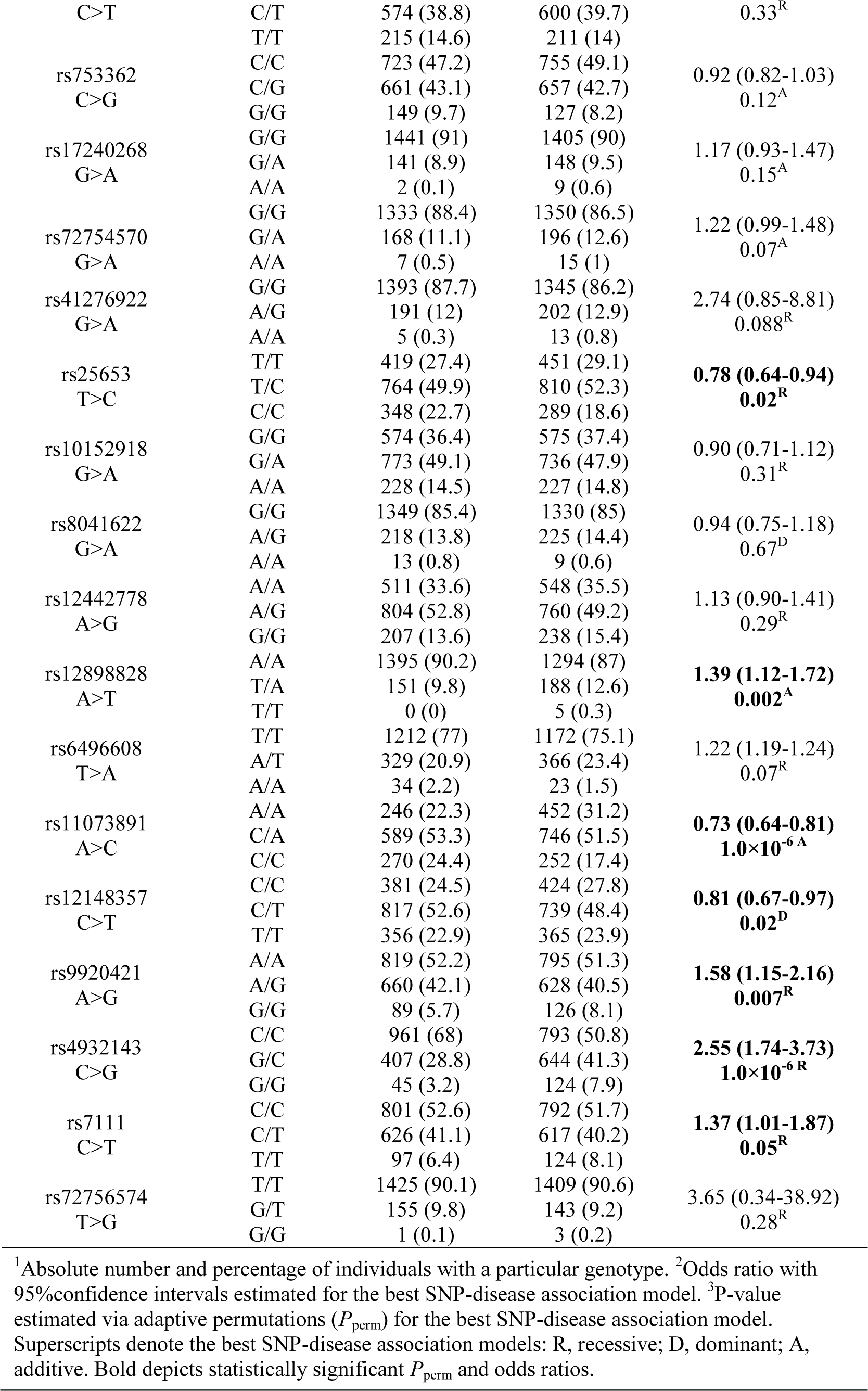
Associations of *ANPEP* genotypes with the risk of type 2 diabetes.

### 3.2 The joint effects of the ANPEP polymorphisms on the risk of T2D

The joint effects of *ANPEP* gene polymorphisms on the risk of type 2 diabetes were assessed through haplotype and diplotype association analyses. The haplotypes and their association with T2D risk are presented in Table 3 (associations of *ANPEP* haplotypes with T2D in sex-stratified groups are shown in Supplementary Table D). Figure 1 displays the linkage disequilibrium map of the *ANPEP* gene, showing four haplotype blocks identified in the gene. As can be seen from Table 3, three haplotypes AATC (block 3), GGT, and GCT (block 4) were strongly (P_perm_<0.0001) associated with susceptibility to type 2 diabetes. Furthermore, two haplotypes such as GGC (block 2) and ATTA (block 3) were associated with the risk of T2D (P_perm_≤0.05).

**Figure 1.**
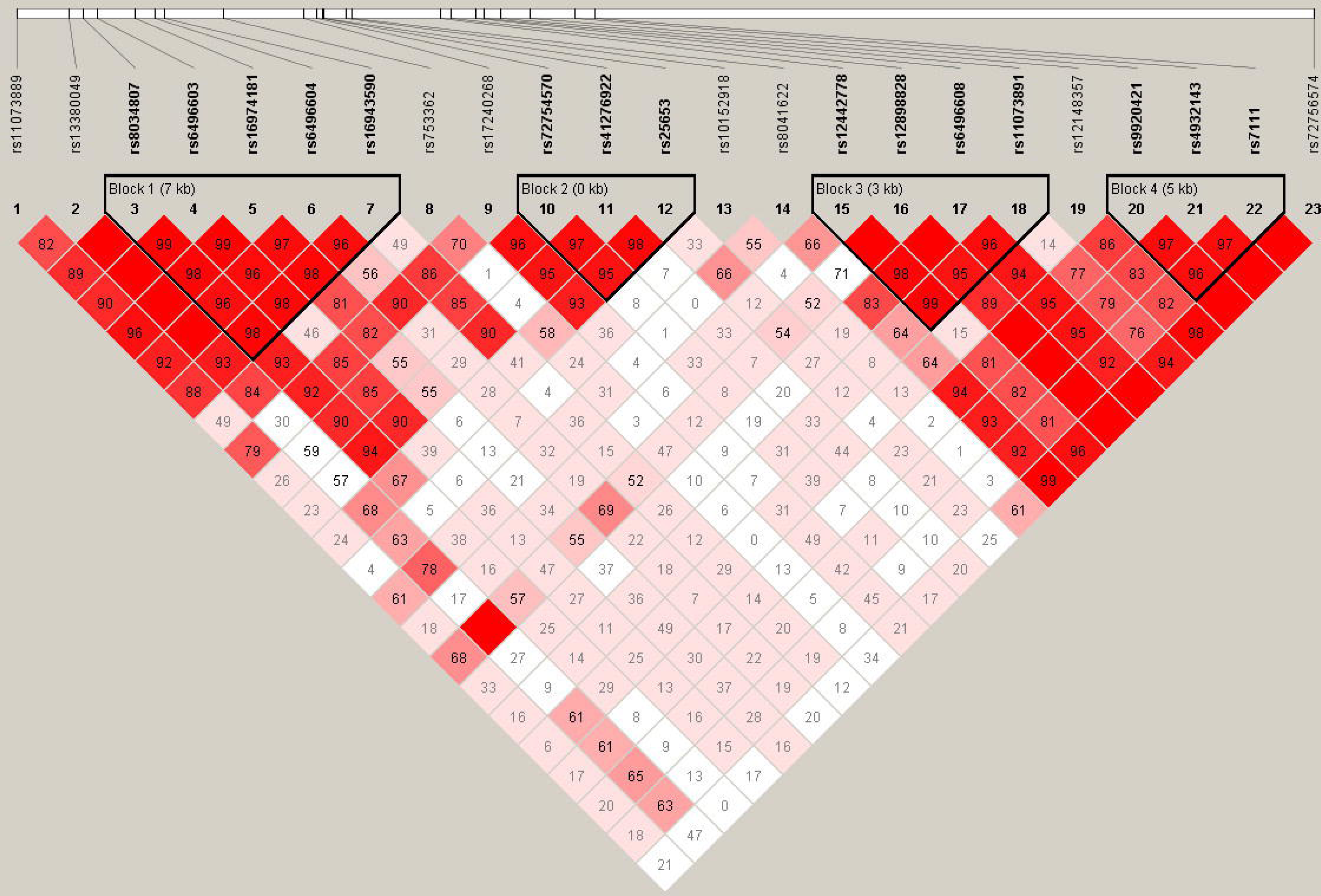
The linkage disequilibrium map of the *ANPEP* gene polymorphisms.

**Table 3.** Haplotypes of the *ANPEP* gene and their associations with T2D risk.

| Haplotype | Haplotype frequencies | | Chi Square | $P_{\text{perm}}$ |
| --- | --- | --- | --- | --- |
|  | T2D patients | Controls |  |  |

| Haplotype block 1<br>rs8034807-rs6496603-rs16974181-rs6496604-rs16943590 |  |  |  |  |
| --- | --- | --- | --- | --- |
| CACAC | 0.490 | 0.494 | 0.13 | 1.00 |
| AGCGT | 0.268 | 0.262 | 0.32 | 1.00 |
| CAGGC | 0.124 | 0.123 | 0.05 | 1.00 |
| CGCGT | 0.066 | 0.070 | 0.31 | 1.00 |
| CGCGC | 0.043 | 0.036 | 2.44 | 0.81 |
| Haplotype block 2<br>rs72754570-rs41276922-rs25653 |  |  |  |  |
| GGT | 0.478 | 0.459 | 2.29 | 0.85 |
| GGC | 0.448 | 0.477 | 5.30 | **0.02** |
| AAT | 0.071 | 0.062 | 2.00 | 0.90 |
| Haplotype block 3<br>rs12442778-rs12898828-rs6496608-rs11073891 |  |  |  |  |
| GATC | 0.400 | 0.408 | 0.42 | 1.00 |
| AATA | 0.389 | 0.382 | 0.33 | 1.00 |
| AAAA | 0.129 | 0.124 | 0.42 | 1.00 |
| ATTA | 0.063 | 0.048 | 7.46 | **0.05** |
| AATC | 0.013 | 0.033 | 28.23 | **$1.0 \times 10^{-7}$** |
| Haplotype block 4<br>rs9920421-rs4932143-rs7111 |  |  |  |  |
| ACC | 0.710 | 0.724 | 1.50 | 0.98 |
| GGT | 0.274 | 0.200 | 47.38 | **$5.9 \times 10^{-12}$** |
| GCT | 0.002 | 0.061 | 174.83 | **$6.5 \times 10^{-40}$** |

The estimation of haplotype frequencies and significance levels for haplotype-disease associations was performed by the Haploview software v.4.2.

Diplotype association analysis allowed for the identification of 136 statistically significant associations (FDR≤0.05) between combinations of *ANPEP* genotypes and an increased (N=78) or decreased (N=58) risk of T2D (all observed associations of diplotypes with T2D risk in the entire and sex-stratified groups are presented in Supplementary Table E). The top-20 diplotypes significantly (FDR-adjusted P≤2.2×10^-8^) associated with the risk of type 2 diabetes are presented in Table 4. The majority of the diplotypes (90 of 136) included the SNP rs4932143. Importantly, *ANPEP* diplotypes such as rs10152918-G/A×rs4931143-G/G (OR 3.35, 95% CI 1.95-5.77, FDR=0.0001), rs25653-T/C×rs4931143-G/G (OR 3.29, 95% CI 1.92-5.66, FDR=0.0002), and rs11073889-C/C×rs4931143-G/G (OR 3.03, 95% CI 1.58-5.81, FDR=0.01) conferred more than a three-fold T2D risk. Diplotype

Four haplotype blocks (bolded frames) were identified within the fifteen SNPs in *ANPEP*. The length of each haplotype block is shown in kilobases, and pairwise linkage disequilibrium (D’) is given for each SNP pair. Empty squares indicate D= 1.0. Linkage disequilibrium map was generated by the Haploview software 4.2

rs9920421-A/G×rs4931143-C/C showed the strongest association with decreased disease risk (OR 0.03, 95% CI 0.01-0.08, FDR=5.7×10^-29^).

**Table 4.**
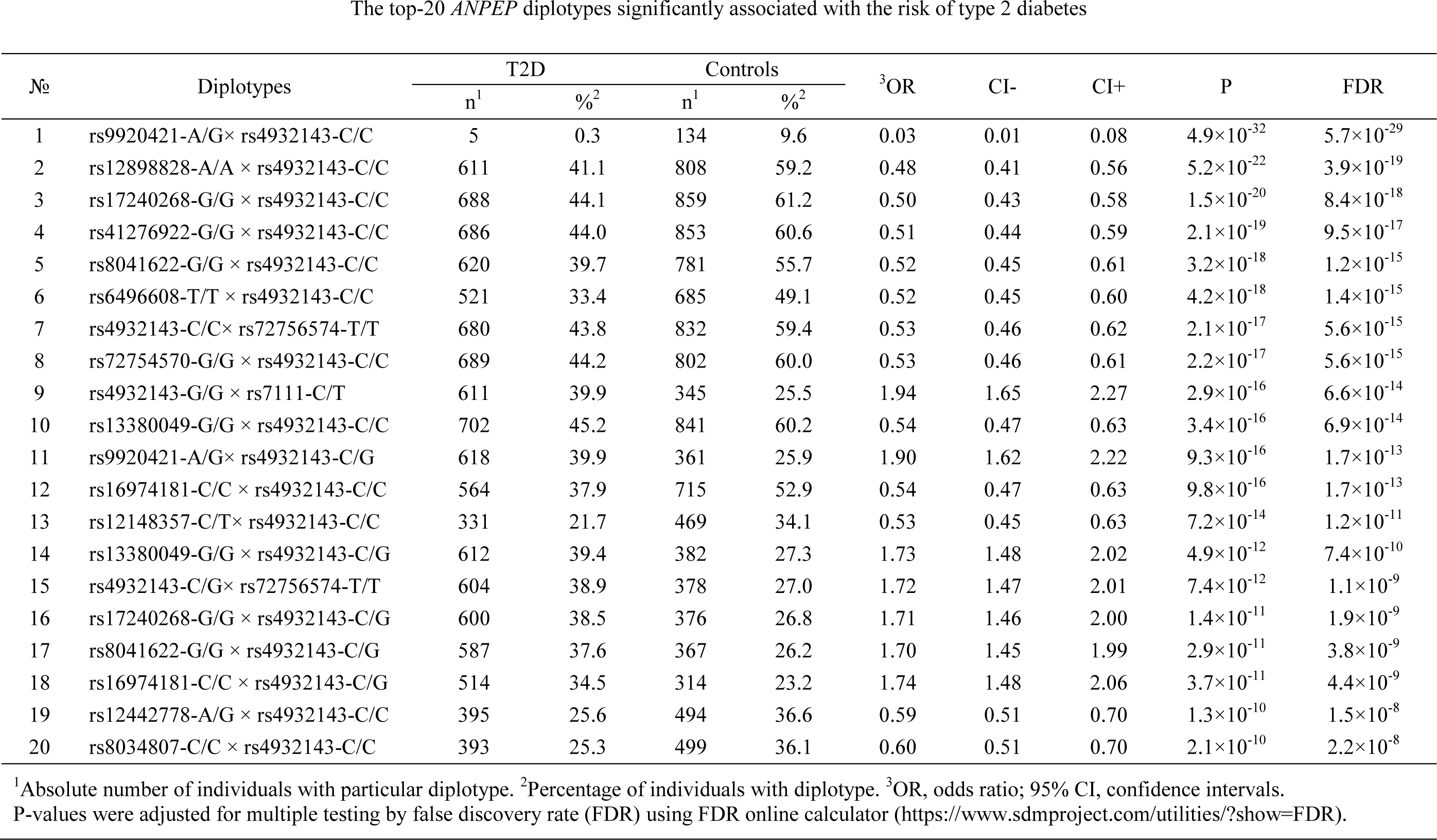
The top-20 *ANPEP* diplotypes significantly associated with the risk of type 2 diabetes.

### 3.3 The link between ANPEP polymorphisms and biochemical parameters in diabetics

The associations between *ANPEP* polymorphisms and biochemical parameters (glucose metabolism, lipid metabolism, redox homeostasis and other biochemical parameters) in the plasma of T2D patients are shown in Supplementary Table F. Figure 2 provides a schematic representation of the observed associations between *ANPEP* polymorphisms and plasma biochemical parameters in patients with type 2 diabetes. The T2D linked SNP rs25653 was associated with higher fasting blood glucose levels (P=0.017), postprandial hyperglycemia in females (P=0.017), and decreased total protein (P=0.008). Polymorphism rs11073891, another variant of *ANPEP* associated with T2D, was correlated with decreased uric acid levels (P=0.03). In females, SNP rs10152918 was positively correlated with total protein (P=0.006) and aspartate aminotransferase (P=0.03) levels. Increased aspartate aminotransferase levels were also associated with the polymorphism rs6496603 (P=0.04). Polymorphism rs17240268 was associated with decreased postprandial hyperglycemia in females (P=0.037) and increased levels of Male and female symbols depict sex-specific SNP-trait associations. Red and blue lines indicate positive and negative associations between SNPs and biochemical parameters, respectively. SNPs associated with the risk of T2D are indicated in red low-density lipoprotein (LDL) cholesterol in males (P=0.017). Decreased LDL levels in the entire group (P=0.04) and increased glutathione concentration in females (P=0.005) correlated with SNP rs72754570. Decreased LDL levels also correlated with rs11073889 in females (P=0.009) and rs12148357 (P=0.04) in the entire group of T2D patients. SNP rs753362 was associated with plasma ROS level in females (P=0.009). Supplementary Table G shows the associations of *ANPEP* haplotypes with biochemical blood plasma parameters of T2D patients. Strong haplotype-trait associations were the subject of great interest. In particular, haplotype CGCGC (haplotype block 1) was strongly associated with increased ROS levels (P<0.0001) and decreased uric acid levels (P<0.0001). Interestingly, haplotype AATC (haplotype block 3), which confers strong protection against type 2 diabetes, was associated with decreased plasma uric acid levels in diabetics (P<0.0001). Meanwhile, haplotype ATTA of the same haplotype block was associated with increased uric acid levels (P<0.0001) and also associated with increased T2D risk. Moreover, haplotype GCT (haplotype block 4), which confers strong protection against disease, was strongly associated with uric acid levels (P<0.0001). In addition, haplotype AATA (haplotype block 3) was associated with decreased uric acid levels (P=0.009).

**Figure 2.**
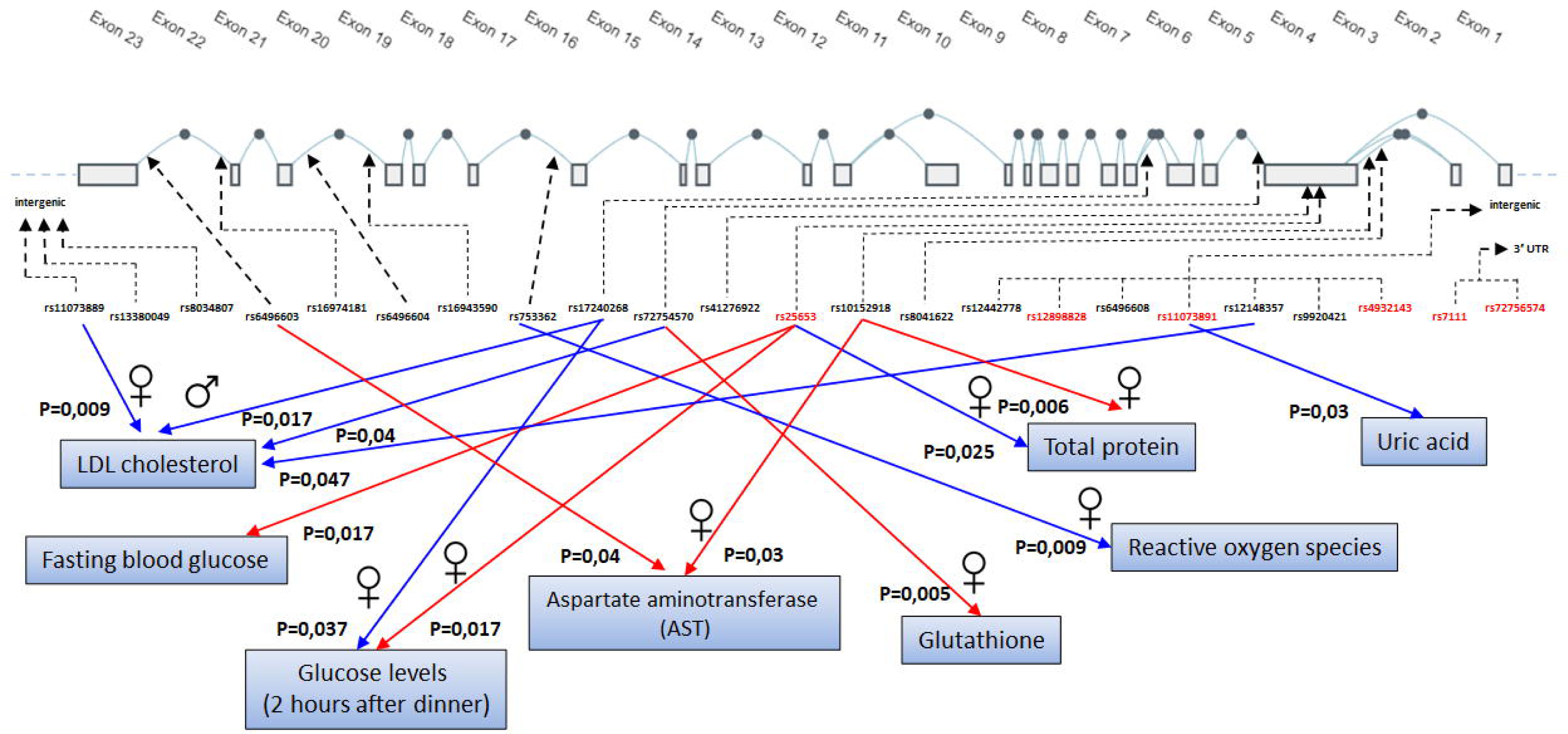
The relationships between *ANPEP* gene polymorphisms and plasma biochemical parameters in diabetic patients.

### 3.4 Replication of the observed SNP-disease associations in independent cohorts

Supplementary Table H shows the associations between the studied *ANPEP* polymorphisms and T2D risk in in 13 cohorts from diverse global populations. The vast majority of SNPs (with the exception of rs25653) including rs12898828, rs11073891, rs12148357, rs9920421, rs4932143, and rs7111, which have been linked to the risk of type 2 diabetes in our population, have been successfully validated in at least one independent population from around the world. Interestingly, ten *ANPEP* polymorphisms, namely, rs11073889, rs13380049, rs6496603, rs6496604, rs753362, rs10152918, rs8041622, rs12442778, rs6496608, and rs72756574, which were not associated with T2D risk in the present study, were found to be associated with disease susceptibility in at least one other population. The present data, despite exhibiting variability in SNP-disease correlations across different populations, unequivocally demonstrate the crucial role that *ANPEP* plays in the development of type 2 diabetes mellitus as a susceptibility gene.

### 3.5 Functional annotation of T2D-associated polymorphisms

Supplementary Table I displays the results of the expression Quantitative Trait Loci (eQTL) analysis for the investigated polymorphisms of the *ANPEP* gene. As per the data provided by the eQTLGen Consortium (https://www.eqtlgen.org/phase1.html), all of the gene variants examined in the current study exhibit an association with increased *ANPEP* expression levels in the blood. As can be seen from Supplementary Table I, the top two T2D-associated SNPs such as rs4932143 and rs11073891 (P_perm_=1.0×10^-6^) were found to associated with expression levels of the *AP3S2* gene in both pancreas and liver. In the blood, the T2D-associated polymorphisms are associated with expression levels of *AP3S2 ARPIN, IDH2, MESP1,* and *U7* genes. Data of the Roadmap Epigenomics project on chromatin states and histone marks in pancreatic islets, pancreas and liver for the *ANPEP* polymorphisms associated with type 2 diabetes mellitus are shown in Supplementary Table J. The region where the polymorphism rs4932143 is located has been shown to exhibit weak enhancer activity in pancreatic islets. Additionally, this region is enriched with active enhancers, which are segments of DNA that interact with transcription factors and play a crucial role in gene expression. The presence of these active enhancers is indicated by the presence of specific histone modifications, such as H3K4me1 and H3K27ac, which are known to be associated with active gene expression. Moreover, the region of rs4932143 also shows primary DNase activity in the pancreas, which suggests that this region is actively transcribed. SNP rs25653, also showed association with T2D, was significantly enriched with regions of active transcription start sites (TSS) and promoter-specific histone modification (H3K4me3) in pancreatic islets, as well as with active, transcribed, and regulatory TSS and histone marks in both the pancreas and liver.

## 4. Discussion

Aminopeptidase N is a type II integral membrane protein that is situated on the plasma membrane as an extracellular enzyme [48,60] with a broad-specificity (there are 151 substrates according to BRENDA (https://www.brenda-enzymes.org). The enzyme is distributed among species and tissues and is abundant in the kidney, small intestine, placenta, and liver [48], with soluble forms found in plasma and urine [61,62]. Besides playing a role in the final digestion of peptides that generated by hydrolysis with gastric and pancreatic proteases, ANPEP is involved in many cellular processes including proliferation, apoptosis, antigen presentation, differentiation, secretion, invasion, angiogenesis, chemotaxis, modulation of bioactive peptide responses and immune functions [60,63–66]. According to the BRENDA (https://www.brenda-enzymes.org, EC3.4.11.2) and KEGG (https://www.genome.jp/kegg/kegg2.html, K11140) databases, ANPEP is a part of glutathione metabolism where the enzyme hydrolyzes dipeptides cysteinyl-glycine (Cys-Gly) and L-cysteinyl-glycine (L-Cys-Gly) generated during the catabolism of glutathione by γ-glutamyltranspeptidase or gamma-glutamyltransferase (GGT) [49,67]. Thus, ANPEP supplies the cell with two key amino acids required for the biosynthesis of glutathione, cysteine, and glycine.

ANPEP has become an object of interest in molecular diabetology owing to the results of genome-wide association studies (GWAS) and transcriptomic analysis of beta cells from pancreatic islets. A GWAS performed in a South Asian population by Kooner et al. was the first to identify *ANPEP* as a possible candidate gene for type 2 diabetes [51]. Two further GWAS studies have identified polymorphisms rs7168849 and rs4932143 located near or in the *ANPEP* gene, respectively, as susceptibility markers for T2D [68,69]. A transcriptome study of pancreatic beta cells from patients with type 2 diabetes showed that *ANPEP* is one of the few genes whose expression was increased in diabetic samples compared to non-diabetic controls [50]. Targeted allelic expression profiling in human islets provided evidence that T2D–associated risk alleles exert detrimental effects by increasing the expression of the *ANPEP* gene [52]. Volkmar et al. revealed that the ANPEP promoter is located in a region that is hypomethylated in the pancreatic islets of patients with T2D [70]. A study by Ligthart et al. showed a tobacco smoking-associated decrease in methylation at CpG island cg23161492, located near the 5′ untranslated region of ANPEP, which is correlated with increased expression of the gene in T2D patients [71]. Collectively, the above data clearly demonstrate the importance of ANPEP in pancreatic islets in the pathogenesis of type 2 diabetes; however, no studies have comprehensively assessed the contribution of *ANPEP* gene polymorphisms to the disease development. Therefore, the present study aimed to explore the joint contribution of functionally significant polymorphisms within the *ANPEP* gene to type 2 diabetes mellitus as well as their relationship with biochemical parameters in patients.

Our study is the first to demonstrate the synergistic effects of 23 *ANPEP* polymorphisms on the risk of type 2 diabetes. Six polymorphisms, rs11073891, rs12898828, rs12148357, rs9920421, rs7111, and rs25653, were found to be associated with type 2 diabetes for the first time. Some SNPs showed sex-dependent effects on disease risk. Several *ANPEP* haplotypes were strongly associated with T2D susceptibility. Diplotype analysis revealed 38 genotype combinations in males and 109 genotype combinations in females that were significantly associated with a predisposition to T2D. A replication analysis of the associations between the studied *ANPEP* gene polymorphisms and T2D susceptibility in 13 populations of the UK Biobank and T2D Knowledge portal confirmed six out of seven associations of *ANPEP* loci with T2D risk established in the Central Russian population. In addition, we found that ten SNPs not linked to T2D in our population were associated with disease risk in at least one of the analyzed independent cohorts. We also established sex-specific associations between *ANPEP* polymorphisms and biochemical parameters in patients with type 2 diabetes, such as fasting blood glucose, postprandial hyperglycemia, total blood protein, glutathione, reactive oxygen species, aspartate aminotransferase, and low-density lipoprotein cholesterol levels. Thus, the present study is the first to investigate and establish the associations of a wide range of functional SNPs of the *ANPEP* gene with the risk of developing type 2 diabetes mellitus, as well as with biochemical and metabolic parameters of patients.

We found that T2D-associated polymorphisms are functionally significant variants associated with changes in the expression of the *ANPEP* gene and genes located in close proximity to *ANPEP*, such as *AP3S2*, *ARPIN*, *IDH2*, *MESP1*, and *U7*. In particular, SNP rs11073891 is enriched with the H3K4me2 histone mark that reliably defines the transcription factor binding site [72] and at the same time is linked to lower expression of the *AP3S2* gene in the pancreas. Interestingly, AP3S2 is involved in regulating vesicle transport in several tissues, including adipocytes and pancreatic cells, and is also associated with first-phase insulin secretion [73] and type 2 diabetes mellitus [51,74,75]. SNP rs12148357 is associated with the ATAC-seq mark, which is analogous to the DNase mark, indicates an accessible region of chromatin [76], and is also associated with decreased pancreatic expression of the *AP3S2* gene. In the pancreas, rs25653 is enriched with enhancer (H3K4me1 and H3K27ac) and promoter (H3K4me3) histone marks that regulate cell type-specific gene expression by facilitating the transcription of target genes [77]. Moreover, in the pancreas, rs25653 falls into the DNAse hypersensitive site, which corresponds to the most active regulatory region [78]. In pancreatic islets, rs25653 is associated with H3K4me3 histone mark in the *ANPEP* promoter.

Polymorphisms of the *ANPEP* gene associated with T2D are correlated with the expression of other genes, such as *ARPIN, MESP1, IDH2, U7*, and *PEX11A*. ARPIN was enriched with GO terms including biological processes such as Negative Regulation Of Cell Migration (GO:0030336), Negative Regulation Of Cell Motility (GO:2000146), and Negative Regulation Of Cytoskeleton Organization (GO:0051494). Similar to Rac, whose gene polymorphisms are associated with T2D risk [57], ARPIN regulates actin polymerization [79], a process essential for the regulation of insulin secretion in pancreatic beta cells [80]. According to the GWAS catalogue (https://www.ebi.ac.uk/gwas/home), several SNPs located between the *ARPIN* and *AP3S2* genes are associated with the risk of type 2 diabetes. MESP1 (mesoderm posterior protein 1) is a transcription factor that regulates cardiac development, morphogenesis, and differentiation. PEX11A (peroxisomal membrane protein 11A) is a member of the peroxisome pathway, and experimental studies in *Pex11a* knockout mice have shown reduced physical activity, decreased fatty acid β-oxidation, and increased de novo lipogenesis, leading to dyslipidemia and obesity [81]. U7 is a small nuclear RNA required for histone pre-mRNA processing [82]. IDH2, a mitochondrial NADP isocitrate dehydrogenase of the pyruvate dehydrogenase complex, plays a role in intermediary metabolism and energy production [83] and supplies NADPH to glutathione reductase for the regeneration of reduced glutathione, namely the reduction of GSSG (oxidized glutathione) to GSH [84]. In pancreatic beta cells, knockdown of mitochondrial isocitrate dehydrogenase enzyme reduces insulin secretion [85]. It has been recently found [23] that pancreatic beta cells in T2D patients exhibit decreased expression of *IDH2*, which strongly correlates with increased expression of *CTH* (cystathionine gamma-lyase), an enzyme in the transsulfuration pathway. It can be assumed that the correlations between the *ANPEP* polymorphisms and expression levels of *ARPIN, MESP1, U7, PEX11A*, and *IDH2* genes may be attributed to shared enhancers co-regulating the expression of these genes co-localized at the 15q26.1 region. These correlations may reflect the crosstalk between biological processes, such as glutathione and energy metabolism, vesicular transport, and insulin secretion by beta cells.

Undertaking a comprehensive analysis of the existing literature to elucidate the molecular mechanisms that underlie the association between ANPEP and type 2 diabetes represents the primary objective of this study. We were also interested in explaining the reasons for the changes in serum levels and other enzymes involved in the metabolism of amino acids and peptides in type 2 diabetes. Elevated plasma levels of liver enzymes, such as gamma-glutamyltransferase (GGT), alanine aminotransferase (ALT), and aspartate aminotransferase (AST), often indicate inflammation or damage to cells in the liver, and these biochemical changes may accompany non-liver pathologies [86]. A plethora of studies [87–95] have reported increased plasma levels or activity of GGT, ALT, and AST in patients with type 2 diabetes. The causes and mechanisms of changes in these enzymes are the subject of debate, but there is still no consensus on whether they are primary or secondary in the development of diabetes or if they accompany the disease. The same can be said for ANPEP, whose expression level is increased both in the plasma and beta cells of the pancreatic islets in patients with type 2 diabetes [23,50,96]. The main assumption is that increased expression of amino acid and peptide metabolism enzymes is required to provide amino acid precursors for the biosynthesis of glutathione, the deficit of which is considered a primary cause of type 2 diabetes [21,31,35]. Numerous studies (reviewed in [22]) have shown that glutathione deficiency and impaired redox homeostasis are etiologically associated with key changes found in type 2 diabetes, such as oxidative stress, mitochondrial dysfunction, endoplasmic reticulum stress, disruption of the ubiquitin-proteasome system and autophagy, disruption of insulin secretion, beta cell dysfunction, and insulin resistance. The factors responsible for endogenous glutathione deficiency in type 2 diabetes mellitus have recently been reviewed in detail [22]. Glutathione deficiency determines the demand of the host for the amino acids such as cysteine, glutamate and glycine required for GSH production. The only mechanism by which the body can compensate for the endogenous deficiency of glutathione under conditions of insufficient dietary intake of its amino acid precursors is the activation of metabolic pathways that produce these amino acids.

We analyzed the KEGG database (https://www.kegg.jp/kegg/, date of access 20.02.2024) on the pathway maps supplying amino acid precursors for glutathione biosynthesis (Glutathione Metabolism, Cysteine and Methionine Metabolism, Alanine, Aspartate and Glutamate Metabolism, Glycine, Serine, and Threonine Metabolism) and summarized the biochemical reactions catalyzed by the enzymes in Figure 3. It is known that many tissues with a limited capacity to synthesize GSH, such as the pancreas, require amino acid precursors from extracellular glutathione synthesized in the liver (the primary site of glutathione synthesis within the body) to transport and enter the cell [97–99]. The figure 3 depicts the primary pathways responsible for the synthesis of precursor amino acids necessary for the production of glutathione. As shown in Figure 3, several pathways are capable of promoting the production of amino acid precursors for GSH biosynthesis. According to the KEGG database, the enzymes responsible for the formation of glutamate, the first amino acid in the glutathione structure, are primarily alanine aminotransferase and aspartate aminotransferase. In addition, glutamate can be produced in the mitochondria from glutamine in the reaction catalyzed by GLS2 (liver isoform of glutaminase, mitochondrial) and from L-1-pyrroline-5-carboxylic acid catalyzed by ALDH4A1 (delta-1-pyrroline-5-carboxylate dehydrogenase, mitochondrial). ALT and AST are enzymes expressed in the liver at high levels that catalyze the transfer of amino groups to generate products in gluconeogenesis and amino acid metabolism. These enzymes utilize amino acids, such as L-alanine (ALT) and L-aspartate (AST), with 2-oxoglutarate derived from the tricarboxylic acid cycle to produce L-glutamate [100]. Increased plasma levels of these enzymes, regularly detected in type 2 diabetes, have been found to be associated with hyperinsulinemia, decreased insulin sensitivity, and hyperglycemia [101,102]. Moreover, the AST/ALT ratio was found to be negatively associated with type 2 diabetes [103–106]. According to the GWAS catalogue (https://www.ebi.ac.uk/gwas/home), the levels of both ALT and AST in the serum are associated with *ANPEP* polymorphisms. We suppose that the increased levels of the liver enzymes reflect an adaptive mechanism in type 2 diabetes aimed at replenishing glutathione deficiency by activating glutamate production for GSH biosynthesis. Cysteine, the second amino acid in the glutathione structure, is produced from the amino acid methionine in the transsulfuration pathway, which converts homocysteine to cysteine. Cystathionine β-synthase (CBS) is a major enzyme in the transsulfuration pathway that catalyzes the condensation of serine and homocysteine to cystathionine through the formation of hydrogen sulfide. Interestingly, the livers of streptozotocin-induced diabetic rats exhibit a marked increase in CBS mRNA levels [107]. A recent experimental study in mice showed that knockout of the CBS gene impairs stimulated insulin secretion and sensitivity [108], suggesting the importance of the transsulfuration pathway in insulin secretion and overall glucose homeostasis. Cystathionine gamma-lyase (CTH) is an enzyme that catalyzes the last step in the transsulfuration pathway by cleaving the L,L-cystathionine molecule into L-cysteine, part of which is utilized for the biosynthesis of glutathione [109]. Our recent study showed up-regulated expression of the *CTH* gene in pancreatic β-cells of T2D patients compared to non-diabetic controls, and the levels of *CTH* were correlated with genes regulating glutathione metabolism and protein folding [23]. Thus, the increased levels of enzymes in the transsulfuration pathway may be of great importance in providing the cell with cysteine for glutathione synthesis. Glycine, the third acid in glutathione, is generated from serine, alanine, threonine, choline, and hydroxyproline through inter-organ metabolism, principally in the liver and kidneys [110]. In particular, it can be formed from alanine and glyoxylate in a reaction catalyzed by the enzyme alanine-glyoxylate aminotransferase (AGXT), serine with the participation of serine hydroxymethyltransferase (SHMT), threonine, and sarcosine (Figure 3). The last reaction is catalyzed by sarcosine dehydrogenase (SARDH) and peroxisomal sarcosine oxidase (PIPOX). According to the TWAS hub database (http://twas-hub.org), increased expression of *SARDH* in a variety of human tissues is associated with the diabetic phenotype and was found to be up-regulated (alongside with *Cth*) in the liver of spontaneous diabetic gerbils [111]. Notably, sarcosine is produced by dimethylglycine-dehydrogenase from dimethylglycine, the deficiency of which is observed in type 2 diabetes [112]. Thus, several lines of evidence support the hypothesis that the up-regulation of enzymes producing amino acids such as glutamate, cysteine, and glycine is an adaptive mechanism directed at replenishing glutathione deficiency in type 2 diabetes mellitus.

**Figure 3.**
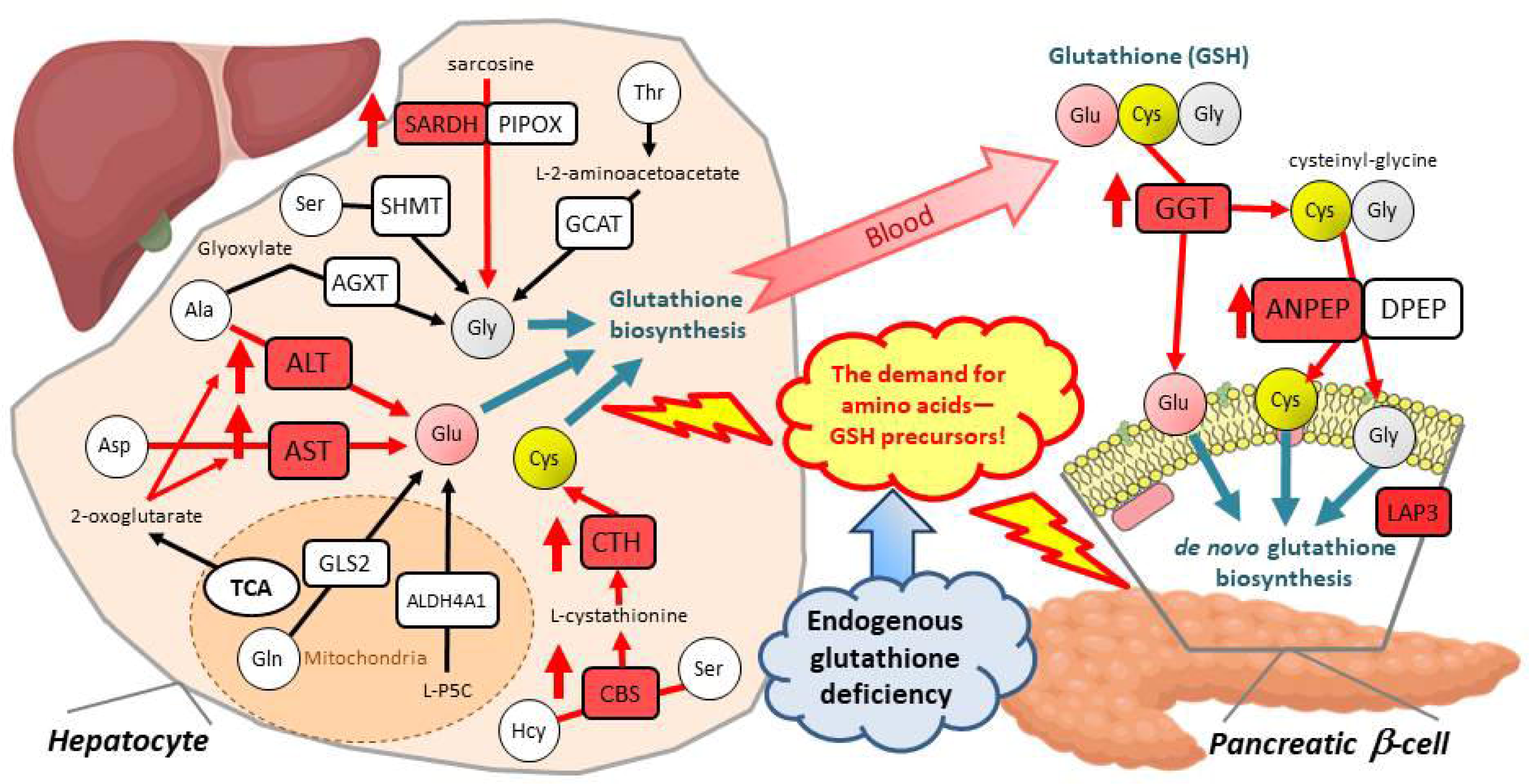
The potential pathways capable of producing amino acids for replenishing glutathione deficiency in type 2 diabetes. ANPEP, membrane alanyl aminopeptidase; GGT, gamma-glutamyltransferase; LAP3, cytosol aminopeptidase; DPEP, dipeptidase; ALT, alanine aminotransferase; AST, alanine transaminase; SARDH, sarcosine dehydrogenase; PIPOX, peroxisomal sarcosine oxidase; GCAT, mitochondrial 2-amino-3-ketobutyrate coenzyme A ligase; SHMT, serine hydroxymethyltransferase; AGXT, alanine-glyoxylate aminotransferase; GLS2, liver isoform of glutaminase (mitochondrial); ALDH4A1, Delta-1-pyrroline-5-carboxylate dehydrogenase (mitochondrial); CTH, cystathionine gamma-lyase; CBS, cystathionine β-synthase; TCA, Tricarboxylic acid cycle; L-P5C, L-1-Pyrroline-5-carboxylic acid. A detailed description of the scheme is presented in the main text.

It is well known that glutathione synthesized in the liver is exported to cells and organs through the blood. Since glutathione cannot enter the cell, membrane-associated enzymes such as gamma-glutamyltransferase, dipeptidase, and alanyl aminopeptidase catabolize glutathione to amino acids, which are then transported into the cell for de novo synthesis of glutathione. According to the UniProt database (https://www.uniprot.org/), gamma-glutamyltransferase catalyzes the transfer of gamma-glutamyl functional groups from molecules such as glutathione, glutathione conjugates, and some leukotrienes to an acceptor that may be amino acids, peptides, or water. The metabolism of glutathione by GGT releases free glutamate and the dipeptide cysteinyl-glycine, which are hydrolyzed to cysteine and glycine by dipeptidases (DPEP) and ANPEP and transported into the cell. As mentioned above, increased levels of both GGT and ANPEP have been found in type 2 diabetes mellitus in numerous studies, whereas no study has investigated the expression levels of dipeptidases in type 2 diabetes. LAP3 (leucine aminopeptidase 3) is a cytosolic metallopeptidase that displays a specific Cys-Gly hydrolyzing activity that produces cysteine and glycine from Cys-Gly-S conjugates [113]. No studies have investigated the levels of LAP3 in T2D. However, a recent study [114] showed that increased LAP3 in the rat liver and serum, as well as in humans, is associated with nonalcoholic fatty liver disease (NAFLD), a condition that regularly co-exists with type 2 diabetes [115]. Additionally, serum LAP3 levels were positively correlated with GGT and fasting blood glucose levels [114]. Moreover, according to the TWAS hub database (http://twas-hub.org), increased levels of LAP3 in blood and various tissue models are associated with the risk of type 2 diabetes mellitus. In the NAFLD patients, ANPEP was found to be strongly correlated with other liver enzymes, such as AST, GGT and alkaline phosphatase [116]. Notably, changes in the levels of enzymes such as GGT, AST, and ALT coexist in type 2 diabetes mellitus and are usually detected before the clinical manifestation of the disease, indicating that these changes are primary; moreover, Mendelian randomization studies did not support a causal relationship between GGT and the risk of type 2 diabetes [117,118]. Similar results were found for the enzymes AST and ALT [119]. In addition, the relationship between ANPEP expression and plasma albumin levels was found to be positively correlated [60], which suggests that co-expression of these two factors may be advantageous in conditions of glutathione deficiency. In such circumstances, ANPEP utilizes the total protein pool, primarily represented by albumin, as a source of peptides and amino acids necessary for the synthesis of GSH. Thus, we hypothesize that the amino acid and peptide metabolism enzymes discussed above are host-upregulated to replenish the glutathione deficiency existing in type 2 diabetes. This hypothesis is supported by the fact that patients with type 2 diabetes exhibit an endogenous deficiency of two amino acids, cysteine [31,120,121] and glycine [122–125]. These amino acids are consequently utilized in two-step reactions of glutathione biosynthesis catalyzed by glutamate cysteine ligase and glutathione synthetase. Notably, deficiency in these amino acids precedes the development of the disease, suggesting the primary nature of the findings. In contrast, the serum levels of glutamate, the third aminoacid in the structure of GSH, were found to be increased in T2D patients [126–128]. It cannot be ruled out that the excessive formation of glutamate in individuals with T2D is an adaptive response to glutathione deficiency and is caused by increased expression of the liver enzymes AST and ALT.

Our study has some limitations. The current study concentrated on a broad spectrum of functionally significant common *ANPEP* polymorphisms; however, it is not possible to rule out the existence of genetic variants, including those that are rare, which could affect an individual’s disease susceptibility independently of the investigated SNPs. Given that polymorphisms associated with T2D are associated not only with the expression of the ANPEP gene (eQTL) but also with other genes, such as *MESP1, PEX11A, AP3S2, PLIN1, WDR93, ARPIN*, *IDH2*, and *U7*, further research, such as that conducted previously [52], should focus on functional studies assessing the phenotypic effects of these SNPs in pancreatic beta cells to identify causative genes. Moreover, further studies in other populations are needed to replicate the observed SNP-disease associations. However, given the heterogeneity of the molecular mechanisms underlying the pathogenesis of type 2 diabetes in different populations worldwide [129], interethnic differences in *ANPEP* polymorphisms associated with disease risk should be expected. Therefore, it is expected that single nucleotide polymorphisms may exhibit weak or moderate phenotypic effects that cannot be replicated in separate populations due to their genetic heterogeneity in minor allele frequencies and linkage disequilibrium between the loci. The limited number of individuals examined for the biochemical variables did not enable us to produce more precise estimates of the connections between *ANPEP* gene variations and both redox homeostasis and intermediate disease phenotypes, such as fasting blood glucose and HbA1c levels. Regrettably, the absence of biochemical data from patients, particularly their plasma levels of ANPEP and GGT, impeded our capacity to analyze molecular-biochemical correlations and provide empirical evidence supporting the hypothesis that ANPEP is associated with glutathione deficiency in individuals with type 2 diabetes. Although our findings support that ANPEP plays an important role in the development of type 2 diabetes, further studies are needed to fully understand the molecular and biochemical mechanisms through which this gene contributes to disease pathogenesis.

The present study generated a hypothesis regarding the role of ANPEP in supplying cells with precursor amino acids required for replenishing glutathione deficiency, which appears to be the primary cause of the development and progression of type 2 diabetes mellitus [21]. In turn, polymorphisms of the *ANPEP* gene are associated with susceptibility to type 2 diabetes mellitus and determine inter-individual differences in gene expression, thereby influencing the efficiency of the final stage of extracellular GSH catabolism through the hydrolysis of cysteinyl-glycine derived from the GGT reaction and providing the cells with key amino acids for de novo synthesis of glutathione. The research conducted on the *ANPEP* gene has not only provided insight into its relationship with type 2 diabetes but has also helped to explain the nature and shared roots of the increased levels of GGT, AST, and ALT enzymes in diabetes. The hypothesis of endogenous deficiency of glutathione in T2D is appealing because it explains both co-existence and shared roots of many disease-associated abnormalities such as mitochondrial dysfunction, increased production of reactive oxygen species, oxidative stress, increased levels of ANPEP, GGT and other liver enzymes, impaired protein folding in the endoplasmic reticulum (ER), ER stress and unfolded protein response regularly observed in diabetics [57].

The traditional explanation of the mechanisms of type 2 diabetes mellitus is based on the initial hypothesis that chronic overnutrition and glucosolipotoxicity are primarily involved in the development of pancreatic beta cell dysfunction and apoptosis [15–20]. However, this hypothesis does not provide an idea of the factors that initiate the cascade of pathological changes in pancreatic β-cells, ultimately leading to apoptosis. We believe that hyperglycemia is a consequence of pancreatic islet cell apoptosis, which is induced by the unfolded protein response pathways due to abnormal protein folding in the endoplasmic reticulum, a condition primarily initiated by disturbances in redox homeostasis and an endogenous deficit of glutathione. Notably, in light of the COVID-19 pandemic, it is important to note that glutathione deficiency was proven by multiple studies as the most likely cause of serious manifestations and death in coronavirus disease [130–135]; thus, it becomes clear why patients with type 2 diabetes mellitus are the most at risk in this group. It is crucial to gain a comprehensive understanding of the relationship between *ANPEP* gene variations, glutathione synthesis, and type 2 diabetes in order to explore potential avenues for correcting disrupted redox homeostasis in patients and to identify scientifically sound interventions for disease prevention and treatment.

## CRediT authorship contribution statement

**Yaroslava Korvyakova**: Investigation, Methodology, Writing – original draft. **Iuliia Azarova**: Resources, Investigation, Formal analysis, Writing – original draft, Funding acquisition. **Elena Klyosova**: Investigation, Methodology, Funding acquisition. **Maria Postnikova**: Investigation, Methodology. **Victor Makarenko**: Software, Methodology, Formal analysis. **Olga Bushueva**: Investigation, Methodology. **Maria Solodilova**: Formal analysis, Validation, Funding acquisition. **Alexey Polonikov**: Conceptualization, Supervision, Investigation, Writing – review & editing, Visualization, Project administration and Funding acquisition.

## Data Availability

All data produced in the present study are available upon reasonable request to the authors

## Data availability

Data supporting reported results are available upon request.

## Acknowledgments

This research was funded by the Russian Science Foundation under Grant no. 20-15-00227. We thank all patients with type 2 diabetes and stuff of the Kursk Emergency Hospital.

## Supplementary data

**Supplementary Table A:** Single nucleotide polymorphisms of the *ANPEP* gene selected for the study

**Supplementary Table B:** A summary of associations between *ANPEP* gene polymorphisms and T2D risk in the sex-stratified groups

**Supplementary Table C:** Associations of *ANPEP* genotypes with the risk of T2D in the sex-stratified groups

**Supplementary Table D:** Haplotypes of the *ANPEP* gene and their associations with T2D risk in the sex-stratified groups

**Supplementary Тable E:** *ANPEP* diplotypes significantly associated with the risk of type 2 diabetes

**Supplementary Table F:** The relationships between *ANPEP* gene polymorphisms and plasma biochemical parameters in diabetic patients

**Supplementary Table G:** The associations of ANPEP haplotypes with biochemical blood plasma parameters of T2D patients

**Supplementary Table H:** A replication analysis for associations between *ANPEP* gene polymorphisms and the risk of type 2 diabetes in independent cohorts

**Supplementary Table I:** eQTL (expression Quantitative Trait Loci) analysis of the studied polymorphisms of the *ANPEP* gene

**Supplementary Table J:** Chromatin states and histone marks in pancreatic islets, pancreas and liver for the *ANPEP* polymorphisms associated with type 2 diabetes mellitus.

