## Supplemental Table A for "The link between the *ANPEP* gene and type 2 diabetes mellitus may be mediated by the disruption of glutathione metabolism and redox homeostasis"

**Supplementary Table A**

Single nucleotide polymorphisms of the *ANPEP* gene selected for the study

| SNP ID | Location | Alleles | Ancestral | MAF | SNP type |
| --- | --- | --- | --- | --- | --- |
| rs11073889 | 15:89778765 | C/A/T | T | 0.42 (C) | intergenic variant |
| rs13380049 | 15:89783507 | G/C | G | 0.12 (C) | regulatory region variant |
| rs8034807 | 15:89784754 | C/A | C | 0.25 (A) | regulatory region variant |
| rs6496603 | 15:89785961 | A/G | A | 0.39 (G) | intron variant |
| rs16974181 | 15:89789408 | C/A/G | G | 0.19 (G) | intron variant |
| rs6496604 | 15:89791246 | A/G | G | 0.41 (A) | intron variant |
| rs16943590 | 15:89792020 | C/G/T | C | 0.31 (T) | intron variant |
| rs753362 | 15:89797303 | G/A/C | G | 0.36 (G) | intron variant |
| rs17240268 | 15:89804583 | G/A | G | 0.07 (A) | missense variant |
| rs72754570 | 15:89805748 | G/A/T | A | 0.08 (A) | intron variant |
| rs41276922 | 15:89806230 | G/A/T | G | 0.04 (A) | missense variant |
| rs25653 | 15:89806327 | C/T | T | 0.42 (C) | missense variant |
| rs10152918 | 15:89808343 | G/A/C | A | 0.40 (A) | intron variant |
| rs8041622 | 15:89808838 | G/A | G | 0.10 (A) | intron variant |
| rs12442778 | 15:89816806 | A/C/G | A | 0.38 (G) | intergenic variant |
| rs12898828 | 15:89817721 | A/T | A | 0.03 (T) | intergenic variant |
| rs6496608 | 15:89819988 | T/A | T | 0.13 (A) | intergenic variant |
| rs11073891 | 15:89820763 | A/C | A | 0.33 (C) | intergenic variant |
| rs12148357 | 15:89822190 | C/G/T | T | 0.47 (C) | intergenic variant |
| rs9920421 | 15:89824835 | G/A/C | G | 0.30 (G) | regulatory region variant |
| rs4932143 | 15:89828835 | G/A/C | C | 0.29 (G) | intergenic variant |
| rs7111 | 15:89830641 | T/C | C | 0.27 (T) | 3 prime UTR variant |
| rs72756574 | 15:89895169 | T/G | T | 0.03 (G) | 3 prime UTR variant |
