## Supplemental Table B for "The link between the *ANPEP* gene and type 2 diabetes mellitus may be mediated by the disruption of glutathione metabolism and redox homeostasis"

**Supplementary Table B**

A summary of associations between *ANPEP* gene polymorphisms and T2D risk in the sex-stratified groups

| SNP ID | Minor Allele | Permutation p-values estimated for tested genetic models of SNP–disease associations^1^ | | | |
| --- | --- | --- | --- | --- | --- |
|  |  | Additive | Allelic | Dominant | Recessive |
| Males | | | | | |
| rs11073889 | C | 0.14 | 0.09 | 0.17 | 0.33 |
| rs13380049 | C | 0.86 | 1.00 | 0.52 | 1.00 |
| rs8034807 | A | 0.73 | 0.73 | 1.00 | 0.86 |
| rs6496603 | G | 0.19 | 0.18 | 0.17 | 0.31 |
| rs16974181 | G | 1.00 | 1.00 | 1.00 | 1.00 |
| rs6496604 | A | 0.67 | 0.56 | 0.73 | 0.52 |
| rs16943590 | T | 0.52 | 0.93 | 1.00 | 0.21 |
| rs753362 | G | 0.86 | 0.59 | 0.67 | 0.78 |
| rs17240268 | A | 0.86 | 1.00 | 0.46 | 1.00 |
| rs72754570 | A | 0.86 | 0.86 | 0.86 | 0.67 |
| rs41276922 | A | 0.86 | 0.86 | 0.78 | 0.56 |
| rs25653 | C | 0.22 | 0.23 | 0.86 | **0.02** |
| rs10152918 | A | 0.73 | 1.00 | 0.38 | 0.86 |
| rs8041622 | A | 0.59 | 0.40 | 1.00 | 0.63 |
| rs12442778 | G | 0.78 | 0.86 | 0.59 | 0.86 |
| rs12898828 | T | 0.86 | 0.20 | 0.37 | NA |
| rs6496608 | A | 0.86 | 0.39 | 0.41 | 1.00 |
| rs11073891 | C | **0.001** | **0.0002** | **0.002** | **0.02** |
| rs12148357 | T | 1.00 | 1.00 | 0.37 | 0.46 |
| rs9920421 | G | 0.86 | 0.86 | 0.48 | 0.52 |
| rs4932143 | G | **0.00001** | **0.000002** | **0.00003** | **0.04** |
| rs7111 | T | 1.00 | 0.86 | 0.78 | 0.63 |
| rs72756574 | G | 0.64 | 0.86 | 1.00 | NA |
| Females | | | | | |
| rs11073889 | C | 0.52 | 1.00 | 0.20 | 0.86 |
| rs13380049 | C | 0.16 | 0.06 | 0.52 | **0.000001** |
| rs8034807 | A | 0.42 | 0.16 | 0.19 | 0.52 |
| rs6496603 | G | 0.86 | 1.00 | 0.48 | 0.15 |
| rs16974181 | G | 0.63 | 0.78 | 0.86 | 0.64 |
| rs6496604 | A | 0.78 | 1.00 | 0.59 | 0.56 |
| rs16943590 | T | 0.86 | 0.86 | 0.64 | 0.06 |
| rs753362 | G | 0.27 | 0.37 | 0.48 | 0.63 |
| rs17240268 | A | 0.11 | 0.06 | 0.13 | NA |
| rs72754570 | A | 0.12 | **0.04** | 0.16 | 0.23 |
| rs41276922 | A | 0.40 | 0.15 | 0.86 | 0.26 |
| rs25653 | C | 0.56 | 0.06 | 1.00 | 0.28 |
| rs10152918 | A | 0.56 | 0.63 | 1.00 | 0.07 |
| rs8041622 | A | 0.28 | 0.73 | 0.26 | 0.78 |
| rs12442778 | G | 0.86 | 1.00 | 1.00 | 0.45 |
| rs12898828 | T | 0.18 | **0.01** | 0.21 | NA |
| rs6496608 | A | 0.37 | 0.86 | 0.78 | **0.01** |
| rs11073891 | C | **0.01** | **0.00001** | 0.06 | **0.03** |
| rs12148357 | T | 0.38 | 0.27 | 0.06 | 0.67 |
| rs9920421 | G | 0.08 | **0.03** | 0.52 | **0.01** |
| rs4932143 | G | **0.000001** | **0.000001** | **0.000001** | **0.00001** |
| rs7111 | T | 0.12 | 0.12 | 0.50 | 0.04 |
| rs72756574 | G | 0.86 | 0.42 | 0.86 | 1.00 |
| ^1^ Calculated by the PLINK software, v.1.9. Bold means statistically significant p-values (P_perm_). | | | | | |
