## Supplemental Table C for "The link between the *ANPEP* gene and type 2 diabetes mellitus may be mediated by the disruption of glutathione metabolism and redox homeostasis"

**Supplementary Table C**

Associations of *ANPEP* genotypes with the risk of T2D in the sex-stratified groups

| SNP ID | Genotype | Genotype frequencies | | | | OR^2^, 95% CI, *P*_perm_^3^ | |
| --- | --- | --- | --- | --- | --- | --- | --- |
|  |  | Males | | Females | | Males | Females |
|  |  | Healthy controls  n (%)^1^ | Patients with T2D  n (%)^1^ | Healthy controls  n (%)^1^ | Patients with T2D  n (%)^1^ |  |  |
| rs11073889  T>C | T/T | 145 (24.7) | 167 (29.2) | 258 (26.6) | 249 (25.6) | 0.87  0,74-1,02  0.09^A^ | 1.18  0.92-1.51  0.20^D^ |
|  | T/C | 301 (51.4) | 281 (49.2) | 487 (50.2) | 507 (52.2) |  |  |
|  | C/C | 140 (23.9) | 123 (21.5) | 225 (23.2) | 216 (22.2) |  |  |
| rs13380049  G>C | G/G | 539 (91.4) | 535 (91.5) | 896 (90.7) | 899 (92.6) | 1.10  0.71-1.71  0.52^D^ | 0.11  0.01-1.16  1.07×10^-6 R^ |
|  | G/C | 50 (8.5) | 48 (8.2) | 88 (8.9) | 71 (7.3) |  |  |
|  | C/C | 1 (0.2) | 2 (0.3) | 4 (0.4) | 1 (0.1) |  |  |
| rs8034807  C>A | C/C | 298 (50.7) | 299 (50.9) | 556 (57.1) | 517 (53.2) | 0.98  0.80-1.18  0.73^A^ | 1.10  0.96-1.27  0.16^A^ |
|  | C/A | 238 (40.5) | 245 (41.7) | 349 (35.8) | 386 (39.8) |  |  |
|  | A/A | 52 (8.8) | 43 (7.3) | 69 (7.1) | 68 (7) |  |  |
| rs6496603  A>G | A/A | 226 (38.3) | 206 (35.3) | 399 (41.1) | 380 (39) | 1.18  0,91-1,52  0.17^D^ | 0.80  0.59-1.09  0.15^R^ |
|  | A/G | 279 (47.3) | 278 (47.7) | 427 (44) | 467 (48) |  |  |
|  | G/G | 85 (14.4) | 99 (17) | 145 (14.9) | 127 (13) |  |  |
| rs16974181  C>G | C/C | 458 (79.5) | 449 (79.3) | 747 (78.6) | 735 (79.5) | 0.99  0,73-1,35  1.0^D^ | 0.95  0.75-1.21  0.56^A^ |
|  | G/C | 112 (19.4) | 108 (19.1) | 185 (19.5) | 171 (18.5) |  |  |
|  | G/G | 6 (1) | 9 (1.6) | 18 (1.9) | 19 (2) |  |  |
| rs6496604  A>G | A/A | 129 (24.3) | 148 (27.2) | 252 (27.6) | 240 (27) | 0.89  0,67-1,20  0.52^R^ | 0.90  0.69-1.16  0.56^R^ |
|  | G/A | 261 (49.2) | 261 (48) | 434 (47.5) | 436 (49.1) |  |  |
|  | G/G | 140 (26.4) | 135 (24.8) | 227 (24.9) | 212 (23.9) |  |  |
| rs16943590  C>T | C/C | 239 (42.9) | 255 (44.7) | 450 (48.9) | 444 (47.2) | 1.26  0,88-1,80  0.21^R^ | 0.74  0.54-1.02  0.057^R^ |
|  | C/T | 244 (43.8) | 223 (39.1) | 330 (35.8) | 377 (40.1) |  |  |
|  | T/T | 74 (13.3) | 92 (16.1) | 141 (15.3) | 119 (12.7) |  |  |
| rs753362  C>G | C/C | 263 (45.9) | 282 (48.9) | 460 (47.9) | 473 (49.2) | 0.91  0,76-1,08  0.59^A^ | 0.91  0.77-1.08  0.27^A^ |
|  | C/G | 251 (43.8) | 241 (41.8) | 410 (42.7) | 416 (43.2) |  |  |
|  | G/G | 59 (10.3) | 54 (9.4) | 90 (9.4) | 73 (7.6) |  |  |
| rs17240268  G>A | G/G | 537 (90.7) | 537 (91.2) | 904 (91.1) | 868 (89.2) | 0.89  0,58-1,37  0.46^D^ | 1.30  0.98-1.74  0.064^A^ |
|  | G/A | 53 (8.9) | 49 (8.3) | 88 (8.9) | 99 (10.2) |  |  |
|  | A/A | 2 (0.3) | 3 (0.5) | 0 (0) | 6 (0.6) |  |  |
| rs72754570  G>A | G/G | 498 (88.5) | 519 (88.1) | 835 (88.4) | 831 (85.5) | 1.51  0,39-5,76  0.67^R^ | **1.31**  **1.01-1.68**  **0.04^A^** |
|  | G/A | 61 (10.8) | 63 (10.7) | 107 (11.3) | 133 (13.7) |  |  |
|  | A/A | 4 (0.7) | 7 (1.2) | 3 (0.3) | 8 (0.8) |  |  |
| rs41276922  G>A | G/G | 523 (88.3) | 516 (87.8) | 870 (87.3) | 829 (85.3) | 1.88  0,41-8,57  0.56^R^ | 1.21  0.95-1.54  0.15^A^ |
|  | A/G | 66 (11.2) | 66 (11.2) | 125 (12.5) | 136 (14) |  |  |
|  | A/A | 3 (0.5) | 6 (1) | 2 (0.2) | 7 (0.7) |  |  |
| rs25653  T>C | T/T | 163 (28.5) | 166 (28.7) | 256 (26.7) | 285 (29.3) | **0.71**  **0.52-0.96**  **0.02^R^** | 0.88  0.78-1.00  0.06^A^ |
|  | T/C | 277 (48.4) | 308 (53.3) | 487 (50.8) | 502 (51.6) |  |  |
|  | C/C | 132 (23.1) | 104 (18) | 216 (22.5) | 185 (19) |  |  |
| rs10152918  G>A | G/G | 210 (35.6) | 221 (38.4) | 364 (37) | 354 (36.8) | 0.89  0,69-1,15  0.38^D^ | 0.76  0.56-1.03  0.07^R^ |
|  | G/A | 306 (51.9) | 266 (46.3) | 467 (47.4) | 470 (48.8) |  |  |
|  | A/A | 74 (12.5) | 88 (15.3) | 154 (15.6) | 139 (14.4) |  |  |
| rs8041622  G>A | G/G | 508 (85.7) | 497 (84.4) | 841 (85.2) | 833 (85.4) | 1.11  0,82-1,48  0.40^A^ | 0.84  0.62-1.14  0.26^D^ |
|  | A/G | 80 (13.5) | 86 (14.6) | 138 (14) | 139 (14.3) |  |  |
|  | A/A | 5 (0.8) | 6 (1) | 8 (0.8) | 3 (0.3) |  |  |
| rs12442778  A>G | A/A | 196 (34.1) | 209 (36.2) | 315 (33.3) | 339 (35) | 0.90  0,69-1,17  0.59^D^ | 1.15  0.84-1.56  0.45^R^ |
|  | A/G | 295 (51.3) | 278 (48.1) | 509 (53.8) | 482 (49.8) |  |  |
|  | G/G | 84 (14.6) | 91 (15.7) | 123 (13) | 147 (15.2) |  |  |
| rs12898828  A>T | A/A | 520 (89.7) | 493 (87.3) | 875 (90.6) | 801 (86.9) | 1.27  0,89-1,80  0.20^A^ | **1.47**  **1.11-1.94**  **0.007^A^** |
|  | T/A | 60 (10.3) | 71 (12.6) | 91 (9.4) | 117 (12.7) |  |  |
|  | T/T | 0 (0) | 1 (0.2) | 0 (0) | 4 (0.4) |  |  |
| rs6496608  T>A | T/T | 456 (77.4) | 441 (75) | 756 (76.7) | 731 (75.1) | 1.11  0,87-1,41  0.39^A^ | **0.32**  **0.12-0.82**  **0.01^R^** |
|  | A/T | 120 (20.4) | 134 (22.8) | 209 (21.2) | 232 (23.8) |  |  |
|  | A/A | 13 (2.2) | 13 (2.2) | 21 (2.1) | 10 (1) |  |  |
| rs11073891  A>C | A/A | 89 (21.3) | 169 (31.5) | 157 (22.9) | 283 (31) | **0.71**  **0,59-0,85**  **1.6×10^-4^** | **0.74**  **0.64-0.85**  **1.0×10^-5 A^** |
|  | C/A | 223 (53.4) | 269 (50.2) | 366 (53.3) | 477 (52.2) |  |  |
|  | C/C | 106 (25.4) | 98 (18.3) | 164 (23.9) | 154 (16.9) |  |  |
| rs12148357  C>T | C/C | 144 (24.8) | 157 (27.2) | 237 (24.3) | 267 (28.1) | 0.85  0,64-1,13  0.37^D^ | 0.78  0.61-1.00  0.06^D^ |
|  | C/T | 294 (50.7) | 264 (45.7) | 523 (53.7) | 475 (50) |  |  |
|  | T/T | 142 (24.5) | 157 (27.2) | 214 (22) | 208 (21.9) |  |  |
| rs9920421  A>G | A/A | 309 (52.6) | 318 (54.6) | 510 (52) | 477 (49.3) | 0.91  0,71-1,16  0.48^D^ | **1.84**  **1.19-2.83**  **0.007^R^** |
|  | A/G | 242 (41.2) | 222 (38.1) | 418 (42.6) | 406 (42) |  |  |
|  | G/G | 36 (6.1) | 42 (7.2) | 53 (5.4) | 84 (8.7) |  |  |
| rs4932143  C>G | C/C | 362 (67.9) | 316 (53.8) | 599 (68.1) | 477 (49) | **1.63**  **1.04-1.34**  **2.0×10^-6 A^** | **2.06**  **1.65-2.58**  **1.0×10^-6 D^** |
|  | G/C | 151 (28.3) | 233 (39.7) | 256 (29.1) | 411 (42.2) |  |  |
|  | G/G | 20 (3.8) | 38 (6.5) | 25 (2.8) | 86 (8.8) |  |  |
| rs7111  C>T | C/C | 303 (53) | 316 (55) | 498 (52.3) | 476 (49.6) | 1.21  0.74-1.98  0.63^R^ | **1.56**  **1.02-2.38**  **0.04^R^** |
|  | C/T | 231 (40.4) | 216 (37.6) | 395 (41.5) | 401 (41.8) |  |  |
|  | T/T | 38 (6.6) | 42 (7.3) | 59 (6.2) | 82 (8.6) |  |  |
| rs72756574  T>G | T/T | 539 (91) | 529 (90.7) | 886 (89.6) | 880 (90.5) | 1.11  0.73-1.68  0.64^A^ | 0.92  0.69-1.22  0.42^A^ |
|  | G/T | 53 (8.9) | 53 (9.1) | 102 (10.3) | 90 (9.3) |  |  |
|  | G/G | 0 (0) | 1 (0.2) | 1 (0.1) | 2 (0.2) |  |  |
| ^1^Absolute number and percentage of individuals with a particular genotype. ^2^Odds ratio with 95%confidence intervals estimated for the best SNP-disease association model. ^3^P-value estimated via adaptive permutations (*P*_perm_) for the best SNP-disease association model. Superscripts denote the best SNP-disease association models: R, recessive; D, dominant; A, additive. Bold depicts statistically significant *P*_perm_ and odds ratios. | | | | | | | |
