## Supplemental Table D for "The link between the *ANPEP* gene and type 2 diabetes mellitus may be mediated by the disruption of glutathione metabolism and redox homeostasis"

**Supplementary Table D**

Haplotypes of the *ANPEP* gene and their associations with T2D risk in the sex-stratified groups

| Haplotype | Haplotype frequencies | | Chi Square | *P*_perm_ |
| --- | --- | --- | --- | --- |
|  | T2D patients | Controls |  |  |
| Males | | | | |
| Haplotype block 1  rs8034807-rs6496603-rs16974181-rs6496604-rs16943590 | | | | |
| CACAC | 0.476 | 0.494 | 0.76 | 1.00 |
| AGCGT | 0.277 | 0.281 | 0.04 | 1.00 |
| CAGGC | 0.115 | 0.112 | 0.06 | 1.00 |
| CGCGT | 0.078 | 0.065 | 1.40 | 0.97 |
| CGCGC | 0.045 | 0.029 | 4.51 | 0.24 |
| Haplotype block 2  rs72754570-rs41276922-rs25653 | | | | |
| GGT | 0.485 | 0.464 | 1.08 | 0.99 |
| GGC | 0.446 | 0.473 | 1.70 | 0.95 |
| AAT | 0.064 | 0.061 | 0.10 | 1.00 |
| Haplotype block 3  rs12442778-rs12898828-rs6496608-rs11073891 | | | | |
| GATC | 0.399 | 0.410 | 0.33 | 1.00 |
| AATA | 0.386 | 0.381 | 0.06 | 1.00 |
| AAAA | 0.130 | 0.120 | 0.60 | 1.00 |
| ATTA | 0.061 | 0.048 | 1.98 | 0.93 |
| AATC | 0.015 | 0.032 | 7.38 | 0.06 |
| Haplotype block 4  rs9920421-rs4932143-rs7111 | | | | |
| ACC | 0.729 | 0.720 | 0.19 | 1.00 |
| GGT | 0.250 | 0.198 | 9.19 | **0.02** |
| GCT | 0.006 | 0.062 | 57.15 | **<0.00001** |
| Females | | | | |
| Haplotype block 1  rs8034807-rs6496603-rs16974181-rs6496604-rs16943590 | | | | |
| CACAC | 0.499 | 0.496 | 0.04 | 1.00 |
| AGCGT | 0.262 | 0.252 | 0.58 | 1.00 |
| CAGGC | 0.129 | 0.127 | 0.04 | 1.00 |
| CGCGT | 0.059 | 0.073 | 2.83 | 0.75 |
| CGCGC | 0.042 | 0.040 | 0.12 | 1.00 |
| Haplotype block 2  rs17240268-rs72754570-rs41276922-rs25653 | | | | |
| GGGC | 0.449 | 0.479 | 3.58 | 0.58 |
| GGGT | 0.473 | 0.455 | 1.26 | 0.99 |
| AAAT | 0.053 | 0.042 | 2.65 | 0.81 |
| GAAT | 0.021 | 0.020 | 0.03 | 1.00 |
| Haplotype block 3  rs12442778-rs12898828-rs6496608-rs11073891 | | | | |
| GATC | 0.402 | 0.407 | 0.12 | 1.00 |
| AATA | 0.391 | 0.382 | 0.33 | 1.00 |
| AAAA | 0.128 | 0.127 | 0.01 | 1.00 |
| ATTA | 0.065 | 0.047 | 5.95 | 0.16 |
| AATC | 0.011 | 0.033 | 21.85 | **<0.00001** |
| Haplotype block 4  rs9920421-rs4932143-rs7111-rs72756574 | | | | |
| ACCT | 0.649 | 0.671 | 2.06 | 0.95 |
| GGTT | 0.288 | 0.202 | 38.8 | **<0.00001** |
| ACCG | 0.048 | 0.053 | 0.40 | 1.00 |
| GCTT | 0.0001 | 0.060 | 118.83 | **<0.00001** |
| The estimation of haplotype frequencies and significance levels for haplotype-disease associations was performed by the Haploview software v.4.2. | | | | |
