## Supplemental Table F for "The link between the *ANPEP* gene and type 2 diabetes mellitus may be mediated by the disruption of glutathione metabolism and redox homeostasis"

**Supplementary Table F**

The relationships between *ANPEP* gene polymorphisms and plasma biochemical parameters in diabetic patients

| SNP ID | Genotype | Entire group | | Males | | | Females | | | |
| --- | --- | --- | --- | --- | --- | --- | --- | --- | --- | --- |
|  |  | Ме (Q1; Q3)^1^ | *P (Q)* | Ме (Q1; Q3)^1^ | | *P (Q)* | Ме (Q1; Q3)^1^ | | *P (Q)* | |
| Fasting blood glucose | | | | | | | | | | |
| rs25653  T>C | T/T | 11,90 (9,3; 15,0) | **0,017**  (0.09) | 12,1 (9,52; 15,1) | 0,10  (0.16) | | 11,6 (9,26; 15) | | 0,072  (0.13) | |
|  | T/C | 12,00 (9,6; 15,0) |  | 12,1 (9,68; 15,0) |  |  | 12 (9,6; 15) | |  |  |
|  | C/C | 12,85 (10,0; 15,8) |  | 12,9 (10,1; 15,6) |  |  | 12,8 (10; 15,92) | |  |  |
| Glucose levels (2 hours after dinner | | | | | | | | | | |
| rs17240268  G>A | G/G | 11,9 (9,2; 14,64) | 0,067  (0.13) | 11,25 (9,06; 14) | 0,33  (0.40) | | 12,2 (9,2; 14,9) | | **0,037**  (0.1) | |
|  | G/A | 11,1 (9,59; 12,4) |  | 11,3 (9,29; 14,5) |  |  | 11,1 (9,8; 12,4) | |  |  |
|  | A/A | 13 (11; 14,5) |  | 18,95 (14,5; 23,4) |  |  | 11,0 (7,51; 13) | |  |  |
| rs25653  T>C | T/T | 11,2 (8,9; 14) | 0,09  (0.15) | 11,1 (8,9; 14) | 0,15  (0.22) | | 11,2 (8,9; 14) | | **0,017**  (0.09) | |
|  | T/C | 12,25 (9,4; 14,64) |  | 12,3 (9,29; 14,64) |  |  | 12,2 (9,4; 14,75) | |  |  |
|  | C/C | 12,4 (9; 15) |  | 10,8 (9; 14) |  |  | 12,75 (8,66; 15,2) | |  |  |
| LDL | | | | | | | | | | |
| rs11073889  T>C | T/T | 3,4 (2,5; 4,55) | **0,029**  (0.10) | 2,8 (2,0; 3,83) | 0,56  (0.62) | | 3,8 (2,79; 4,7) | | **0,0094**  (0.09) | |
|  | T/C | 2,99 (2,32; 3,94) |  | 2,69 (2,1; 3,43) |  |  | 3,13 (2,58; 4,22) | |  |  |
|  | C/C | 2,90 (2,48; 3,89) |  | 2,65 (2,4; 2,99) |  |  | 3,2 (2,5; 4,12) | |  |  |
| rs17240268  G>A | G/G | 3,0 (2,4; 4,0) | 0,12  (0.18) | 2,7 (2,1; 3,5) | **0,017**  (0.09) | | 3,2 (2,57; 4,34) | | 0,13  (0.19) | |
|  | G/A | 3,33 (2,8; 4,48) |  | 2,83 (2,8; 3,6) |  |  | 3,88 (2,9; 4,6) | |  |  |
|  | A/A | 4,1 (3,78; 4,74) |  | 4,1 (4,1; 4,1) |  |  | 4,26 (3,78; 4,74) | |  |  |
| rs72754570  G>A | G/G | 2,99 (2,4; 4,0) | 0,098  (0.16) | 2,7 (2,1; 3,5) | **0,041**  (0.01) | | 3,2 (2,58; 4,36) | | **0,045**  (0.11) | |
|  | G/A | 3,25 (2,75; 4,48) |  | 2,8 (2,23; 3,6) |  |  | 3,8 (2,85; 4,65) | |  |  |
|  | A/A | 3,8 (3,7; 3,86) |  | 3,8 (2,83; 4,1) |  |  | 4,07 (3,74; 4,55) | |  |  |
| rs12148357  C>T | C/C | 3,37 (2,52; 4,4) | **0,047**  (0.10) | 2,7 (2,3; 3,94) | 0,24  (0.31) | | 3,6 (2,72; 4,5) | | 0,089  (0.15) | |
|  | C/T | 3,0 (2,4; 4) |  | 2,8 (2,14; 3,42) |  |  | 3,15 (2,6; 4,4) | |  |  |
|  | T/T | 2,88 (2,26; 3,81) |  | 2,6 (1,9; 3,3) |  |  | 3,23 (2,5; 3,9) | |  |  |
| GSSG | | | | | | | | | | |
| rs72754570  G>A | G/G | 1,35 (0,51; 3,75) | **0,0089**  (0.09) | 1,64 (0,71; 3,79) | | 0,35  (0.42) | | 1,31 (0,44; 3,68) | | **0,0053**  (0.09) |
|  | G/A | 2,99 (0,91; 4,21) |  | 3,66 (0,91; 3,92) | |  |  | 2,47 (1,11; 4,21) | |  |
|  | A/A | 1,94 (1,25; 4,26) |  | 1,25 (1,25; 1,25) | |  |  | 3,10 (1,94; 4,26) | |  |
| ROS | | | | | | | | | | |
| rs753362  C>G | C/C | 3,53 (2,47; 4,81) | **0,0094**  (0.09) | 3,35 (2,20; 4,54) | | 0,58  (0.63) | | 3,55 (2,54; 4,89) | | **0,0085**  (0.09) |
|  | C/G | 4,02 (2,83; 5,29) |  | 3,63 (2,52; 5,03) | |  |  | 4,19 (3,05; 5,38) | |  |
|  | G/G | 3,30 (2,82; 4,28) |  | 3,02 (2,35; 3,92) | |  |  | 3,37 (2,82; 4,47) | |  |
| Uric acid | | | | | | | | | | |
| rs11073891  A>C | A/A | 343,65 (279,6; 419,5) | **0,033**  (0.01) | 356,5 (284,2; 429,5) | | 0,29  (0.36) | | 340,2 (270,1; 416,2) | | 0,13  (0.19) |
|  | C/A | 326,4 (254,9; 401,9) |  | 331,9 (256,1; 392,5) | |  |  | 324,6 (253,3; 403,9) | |  |
|  | C/C | 311,0 (248,6; 368,6) |  | 323,4 (258,6; 394,8) | |  |  | 305,3 (248,2; 368,1) | |  |
| Total protein | | | | | | | | | | |
| rs25653  T>C | T/T | 74 (69; 78) | **0,0079**  (0.09) | 75 (71; 79) | | 0,32  (0.39) | | 73,5 (69; 77) | | **0,025**  (0.09) |
|  | T/C | 73 (69; 77) |  | 74 (69; 78) | |  |  | 72 (68; 76) | |  |
|  | C/C | 73 (69; 78) |  | 72,5 (69; 77) | |  |  | 73 (68,5; 78) | |  |
| rs10152918  G>A | G/G | 72,1 (68; 77) | 0,064  (0.13) | 74 (69; 77) | | 0,93  (0.93) | | 72 (68; 77) | | **0,0059**  (0.09) |
|  | G/A | 73 (69; 77) |  | 74 (69; 78) | |  |  | 73 (69; 77) | |  |
|  | A/A | 74 (70; 78) |  | 73 (69; 79) | |  |  | 74 (70; 78) | |  |
| AST | | | | | | | | | | |
| rs6496603  A>G | A/A | 24 (18; 34) | **0,044**  (0.10) | 26 (20; 38) | | 0,29  (0.36) | | 24 (18; 33) | | 0,08  (0.14) |
|  | A/G | 25 (19; 40) |  | 27 (19; 43) | |  |  | 24 (18; 37) | |  |
|  | G/G | 29 (20; 52) |  | 30 (23; 45,5) | |  |  | 29 (18; 54) | |  |
| rs10152918  G>A | G/G | 24 (18; 33,8) | **0,04**  (0.10) | 25 (19; 36) | | 0,72  (0.76) | | 23 (18; 32) | | **0,033**  (0.10) |
|  | G/A | 25 (18; 35) |  | 26 (19; 41) | |  |  | 24 (18; 34) | |  |
|  | A/A | 26 (19; 41) |  | 26,5 (20; 42) | |  |  | 26 (18;40) | |  |
