## Supplemental Table H for "The link between the *ANPEP* gene and type 2 diabetes mellitus may be mediated by the disruption of glutathione metabolism and redox homeostasis"

**Supplementary Table H**

A replication analysis for associations between *ANPEP* gene polymorphisms and the risk of type 2 diabetes in independent cohorts

| SNP | Minor allele | ^1^Central Russia  (present study) | ^2^UK Biobank 1 | ^3^UK Biobank 2 | ^4^DIAGRAM 1000G GWAS | ^4^PAGE 2019 cardiometabolic traits GWAS: trans-ancestry | ^4^T2D TOPMed imputed 2023 GWAS: Hispanic ancestry | ^4^Diabetic Cohort Singapore Prospec-tive Study SEED GWAS | ^4^DIAMANTE (European) T2D GWAS | ^4^T2D 2022 GWAS: South Asian ancestry | ^4^TOPMed T2D whole genome sequence analysis | ^4^BioMe AMP T2D GWAS | ^4^METSIM GWAS | ^4^CAMP GWAS | ^4^China Health and Nutrition Survey 2018 glycemic traits GWAS: East Asian ancestry |
| --- | --- | --- | --- | --- | --- | --- | --- | --- | --- | --- | --- | --- | --- | --- | --- |
| rs11073889  T>C | C | 0.24 | 0.23 | 0.65 | **0.0055** | **0.014** | 0.44 | 0.95 | 0.056 | 0.49 | 0.40 | - | - | - | 0.72 |
| rs13380049  G>C | C | 0.06 | 0.69 | 0.68 | 0.60 | **0.025** | **0.03** | - | 0.76 | - | 0.32 | - | - | - | 0.57 |
| rs8034807  C>A | A | 0.28 | 0.89 | 0.75 | 0.16 | 0.96 | 0.41 | - | 0.32 | 0.77 | 0.27 | - | - | - | 0.45 |
| rs6496603  A>G | G | 0.18 | 0.90 | 0.60 | 0.37 | 0.20 | 0.15 | **0.036** | 0.52 | 0.85 | 0.61 | 0.66 | - | - | 0.73 |
| rs16974181  C>G | G | 0.78 | 0.74 | 0.13 | 0.70 | 0.44 | 0.89 | - | 0.61 | - | 0.58 | - | - | - | - |
| rs6496604  A>G | G | 0.59 | 0.91 | 0.51 | 0.34 | **0.045** | 0.23 | - | 0.34 | 0.21 | 0.67 | - | - | - | 0.76 |
| rs16943590  C>T | T | 0.33 | 0.99 | 0.99 | 0.17 | 0.055 | 0.058 | - | 0.42 | - | 0.27 | - | - | - | 0.62 |
| rs753362  C>G | G | 0.12 | 0.12 | 0.80 | 0.13 | 0.15 | 0.78 | - | **0.01** | - | 0.64 | - | - | - | 0.27 |
| rs17240268  G>A | A | 0.15 | 0.64 | 0.17 | 0.98 | 0.78 | 0.86 | - | 0.72 | 0.17 | 0.65 | 0.21 | 0.61 | 0.20 | - |
| rs72754570  G>A | A | 0.069 | 0.67 | 0.21 | 0.91 | 0.82 | 0.84 | - | 0.69 | 0.21 | 0.55 | - | - | - | - |
| rs41276922  G>A | A | 0.088 | 0.63 | 0.12 | 0.82 | 0.76 | 0.95 | - | 0.59 | - | 0.71 | - | - | - | - |
| rs25653  T>C | C | **0.015** | 0.19 | 0.22 | 0.52 | 0.49 | 0.60 | 0.62 | 0.21 | 0.98 | 0.37 | - | 0.11 | 0.36 | 0.056 |
| rs10152918  G>A | A | 0.31 | 0.17 | 0.38 | 0.32 | 0.78 | 0.67 | 0.34 | 0.17 | 0.45 | 0.55 | - | **0.014** | - | - |
| rs8041622  G>A | A | 0.67 | 0.24 | **0.013** | 0.23 | 0.86 | 0.95 | 0.77 | 0.93 | - | 0.72 | 0.96 | **0.027** | - | - |
| rs12442778  A>G | G | 0.29 | **0.0092** | **0.0032** | 0.69 | 0.66 | 0.12 | - | **3×10^-5^** | **0.037** | 0.30 | - | - | - | 0.06 |
| rs12898828  A>T | T | **0.0021** | 0.32 | **0.029** | 0.15 | 0.58 | 0.54 | - | 0.15 | - | 0.27 | - | - | - | - |
| rs6496608  T>A | A | 0.07 | 0.41 | 0.081 | 0.091 | **0.018** | 0.44 | - | 0.20 | 0.051 | - | - | - | - | - |
| rs11073891  A>C | C | **1×10^-6^** | **0.004** | **0.001** | 0.78 | 0.61 | **0.025** | **0.014** | **4×10^-5^** | **0.026** | 0.68 | 0.76 | 0.76 | **0.0072** | **0.033** |
| rs12148357  C>T | T | **0.023** | 0.47 | **0.027** | **0.0018** | **0.0029** | 0.14 | - | **6×10^-5^** | - | **0.027** | - | - | - | 0.11 |
| rs9920421  A>G | G | **0.0066** | **0.0006** | **1.2×10^-5^** | **0.0013** | **2.5×10^-5^** | 0.87 | - | **6×10^-19^** | **0.0001** | **0.012** | **0.013** | **0.042** | - | 0.11 |
| rs4932143  C>G | G | **1×10^-6^** | **0.0002** | **6.1×10^-6^** | **0.0007** | 0.055 | 0.93 | - | **2×10^-19^** | - | **0.0085** | - | - | - | 0.23 |
| rs7111  C>T | T | **0.046** | **0.0002** | **8.1×10^-6^** | **0.0011** | **0.0001** | 0.91 | 0.36 | **1.5×10^-19^** | **0.0002** | **0.027** | - | - | - | 0.22 |
| rs72756574  T>G | G | 0.42 | 0.32 | 0.91 | 0.73 | 0.94 | 0.95 | - | **0.014** | - | - | - | - | - | - |
| ^1^ Data on the levels of significance of associations of the studied SNPs with the risk of T2D in the population of Central Russia adjusted for sex, age and BMI;  ^2^Data on the levels of significance of associations of the studied SNPs with the risk of T2D in the population UK Biobank (type 2 diabetes mellitus) adjusted for sex, age and BMI (Gene Atlas URL: <http://geneatlas.roslin.ed.ac.uk>);  ^3^Data on the levels of significance of associations of the studied SNPs with the risk of T2D in the population UK Biobank (non-insulin-dependent diabetes mellitus) adjusted for sex, age and BMI (Gene Atlas URL: <http://geneatlas.roslin.ed.ac.uk>);  ^4^Data on the levels of significance of associations of the studied SNPs with the risk of T2D in populations of the Type 2 Diabetes knowledge Portal (type 2 diabetes) adjusted for BMI (URL: <http://www.type2diabetesgenetics.org>). | | | | | | | | | | | | | | | |
