## Supplemental Table I for "The link between the *ANPEP* gene and type 2 diabetes mellitus may be mediated by the disruption of glutathione metabolism and redox homeostasis"

**Supplementary Table I**

eQTL (expression Quantitative Trait Loci) analysis of the studied polymorphisms of the *ANPEP* gene

| SNP ID | | Assessed allele |  | eQTL (expression quantitative trait locus) | | | | | | sQTL (splicing quantitative trait locus) | | | | | | mQTL (methylation quantitative trait locus) in blood^3^ |
| --- | --- | --- | --- | --- | --- | --- | --- | --- | --- | --- | --- | --- | --- | --- | --- | --- |
|  |  |  | Gene | Pancreas^1^ | | Liver^1^ | | Whole blood^2^ | | Pancreas^1^ | | Liver^1^ | | Whole blood ^1^ | |  |
|  |  |  |  | NES | P | NES | P | Z | Q | NES | P | NES | P | NES | P |  |
| rs11073889 | | C | *ANPEP* | -0.06 | 0.24 | -0.02 | 0.68 | 5.0 | **0.002** | - | - | - | - | - | - |  |
|  |  |  | *MESP1* | 0.02 | 0.75 | 0.16 | 0.04 | -27.4 | **5.6×10^-157^** | - | - | - | - | - | - | - |
|  |  |  | *PEX11A* | 0.06 | 0.28 | -0.10 | 0.16 | -11.0 | **4.3×10^-20^** | - | - | - | - | - | - | - |
|  |  |  | *AP3S2* | -0.04 | 0.41 | 0.01 | 0.95 | 7.3 | **3.4×10^-5^** | - | - | - | - | - | - | - |
|  |  |  | *PLIN1* | 0.08 | 0.21 | 0.03 | 0.63 | -5.0 | **0.001** | - | - | - | - | - | - | - |
|  |  |  | *WDR93* | -0.36 (T) | **3.2×10^-6^** | NA | NA | - | - | - | - | - | - | - | - | - |
| rs13380049 | | C | *ANPEP* | 0.04 | 0.70 | 0.02 | 0.86 | 7.43 | **1.4×10^-5^** | - | - | - | - | - | - | 1 |
|  |  |  | *AP3S2* | 0.06 | 0.51 | -0.14 | 0.29 | -11.89 | **1.7×10^-24^** | - | - | - | - | - | - | - |
|  |  |  | *ARPIN* | 0.04 | 0.44 | 0.08 | 0.52 | -9.93 | **3.8×10^-15^** | - | - | - | - | - | - | - |
|  |  |  | *U7* | NA | NA | NA | NA | -5.29 | **0.0004** | - | - | - | - | - | - | - |
| Haplotype block 1 | rs8034807 | A | *ANPEP* | -0.04 | 0.52 | 0.07 | 0.27 | -11.34 | **1.0×10^-21^** | - | - | - | - | - | - | - |
|  |  |  | *MESP1* | 0.13 | 0.03 | -0.02 | 0.76 | 21.80 | **3.2×10^-97^** | - | - | - | - | - | - | - |
|  |  |  | *PLIN1* | -0.05 | 0.46 | 0.04 | 0.57 | -4.76 | **0.005** | - | - | - | - | - | - | - |
|  | rs6496603 | G | *ANPEP* | -0.03 | 0.56 | 0.06 | 0.25 | -9.60 | **9.7×10^-14^** | - | - | - | - | - | - | - |
|  |  |  | *MESP1* | 0.07 | 0.20 | 0.02 | 0.84 | 20.20 | **1.4×10^-82^** | - | - | - | - | - | - | - |
|  |  |  | *AP3S2* | 0.00 | 0.99 | -0.02 | 0.78 | -8.39 | **5.8×10^-9^** | - | - | - | - | - | - | - |
|  |  |  | *PEX11A* | -0.03 | 0.61 | 0.02 | 0.78 | -6.67 | **0.003** | - | - | - | - | - | - | - |
|  | rs16974181 | G | *ANPEP* | 0.02 | 0.20 | 0.02 | 0.02 | - | - | - | - | - | - | 0.32 | - | - |
|  |  |  | *PEX11A* | 0.09 | 0.20 | -0.19 | 0.03 | 23.85 | **1.2×10^-117^** | - | - | - | - | - | - | - |
|  |  |  | *PLIN1* | 0.03 | 0.67 | -0.13 | 0.07 | 9.15 | **7.5×10^-12^** | - | - | - | - | - | - | - |
|  | rs6496604 | G | *ANPEP* | -0.09 | 0.10 | -0.06 | 0.32 | 7.73 (A) | **1.3×10^-6^** | - | - | - | - | - | - | - |
|  |  |  | *MESP1* | 0.05 | 0.40 | 0.14 | 0.07 | -24.06(A) | **8.7×10^-120^** | - | - | - | - | - | - | - |
|  |  |  | *PEX11A* | 0.22 | 0.49 | -0.09 | 0.22 | -11.08 (A) | **1.9×10^-20^** | - | - | - | - | - | - | - |
|  |  |  | *AP3S2* | -0.06 | 0.22 | -0.00 | 0.96 | 9.32 (A) | **1.4×10^-12^** | - | - | - | - | - | - | - |
|  | rs16943590 | T | *ANPEP* | -0.02 | 0.72 | 0.08 | 0.18 | -7.89 | **3.6×10^-7^** | - | - | - | - | - | - | - |
|  |  |  | *MESP1* | 0.10 | 0.09 | 0.02 | 0.79 | 19.26 | **1.5×10^-74^** | - | - | - | - | - | - | - |
|  |  |  | *MESP2* | 0.33 | **0.00005** | 0.00 | 0.98 | - | - | - | - | - | - | - | - | - |
|  |  |  | *AP3S2* | 0.47 | 0.32 | -0.05 | 0.47 | -8.78 | **2.1×10^-10^** | - | - | - | - | - | - | - |
|  |  |  | *PEX11A* | 0.04 | 0.47 | -0.02 | 0.77 | -7.61 | **3.5×10^-6^** | - | - | - | - | - | - | - |
|  |  |  | *PLIN1* | -0.04 | 0.59 | 0.08 | 0.19 | -5.40 | **0.0003** | - | - | - | - | - | - | - |
| rs753362 | | C | *ANPEP* | -0.07 | 0.26 | -0.02 | 0.78 | - | - | - | - | - | - | - | - | - |
| rs17240268 | | A | *ANPEP* | -0.24 | 0.02 | -0.27 | 0.009 | 7.84 | **5.4×10^-7^** | 0.77 | **6.1×10^-16^** | 0.71 | **1.5×10^-6^** | 1.0 | **1.5×10^-44^** | - |
|  |  |  | *PEX11A* | -0.00 | 0.98 | -0.12 | 0.35 | 14.1 | **2.4×10^-37^** | - | - | - | - | - | - | - |
|  |  |  | *PLIN1* | 0.05 | 0.71 | -0.12 | 0.26 | 5.35 | **0.0003** | - | - | - | - | - | - | - |
|  |  |  | *MESP1* | -0.12 | 0.25 | 0.19 | 0.17 | 4.37 | **0.03** | - | - | - | - | - | - | - |
| Haplotype block 2 | rs72754570 | A | *ANPEP* | -0.26 | 0.009 | -0.29 | 0.004 | 7.25 | **5.3×10^-5^** | 0.72 | **4.8×10^-15^** | - | - | 0.95 | - | - |
|  |  |  | *PEX11A* | -0.03 | 0.81 | -0.05 | 0.71 | 13.60 | **4.8×10^-34^** | - | - | - | - | - | - | - |
|  |  |  | *PLIN1* | 0.04 | 0.71 | -0.10 | 0.35 | 5.19 | **0.0006** | - | - | - | - | - | - | - |
|  | rs41276922 | A | *ANPEP* | -0.27 | 0.01 | -0.34 | 0.002 | 6.61 | **0.005** | 0.90 | **1.1×10^-19^** | 0.83 | **1.6×10^-7^** | 1.10 | - | - |
|  |  |  | *PEX11A* | -0.06 | 0.58 | -0.13 | 0.36 | 13.03 | **1.1×10^-30^** | - | - | - | - | - | - | - |
|  |  |  | *PLIN1* | 0.01 | 0.92 | -0.11 | 0.31 | 4.76 | **0.005** | - | - | - | - | - | - | - |
|  | rs25653 | C | *ANPEP* | -0.09 | 0.09 | 0.03 | 0.57 | - | - | - | - | - | - | 0.20 | **1.6×10^-6^** | 4 |
|  |  |  | *MESP1* | -0.04 | 0.43 | -0.02 | 0.78 | -6.4 | **0.02** | - | - | - | - | - | - | - |
|  |  |  | *AP3S2* | 0.02 | 0.66 | 0.04 | 0.61 | -5.6 | **0.0001** | - | - | - | - | - | - | - |
| rs10152918 | | A | *ANPEP* | -0.01 | 0.84 | 0.04 | 0.44 | -12.70 | **7.7×10^-29^** | - | - | - | - | - | - | - |
|  |  |  | *AP3S2* | -0.04 | 0.37 | 0.04 | 0.51 | 5.54 | **0.0001** | - | - | - | - | - | - | - |
| rs8041622 | | A | *ANPEP* | -0.17 | 0.06 | 0.07 | 0.43 | 19.79 | **4.8×10^-79^** | - | - | - | - | - | - | - |
|  |  |  | *AP3S2* | -0.15 | 0.07 | -0.17 | 0.14 | -16.50 | **4.3×10^-53^** | - | - | - | - | - | - | - |
| Haplotype block 3 | rs12442778 | G | *ANPEP* | -0.08 | 0.16 | -0.02 | 0.69 | -106.77 | **4.2×10^-302^** | - | - | - | - | - | - | - |
|  |  |  | *ARPIN* | -0.08 | 0.007 | 0.09 | 0.17 | 17.45 | **4.4×10^-60^** | - | - | - | - | - | - | - |
|  |  |  | *AP3S2* | -0.22 | **0.00001** | -0.26 | 0.00007 | 8.06 | **1.0×10^-7^** | - | - | - | - | - | - | - |
|  |  |  | *IDH2* | -0.03 | 0.34 | -0.02 | 0.62 | 5.81 | **0.00005** | - | - | - | - | - | - | - |
|  |  |  | *MESP1* | 0.01 | 0.82 | -0.02 | 0.76 | 5.30 | **0.0004** | - | - | - | - | - | - | - |
|  | **rs12898828** | T | *ANPEP* | -0.04 | 0.73 | -0.10 | 0.40 | - | **-** | - | - | - | - | - | - | 1 |
|  |  |  | *AP3S2* | -0.24 | 0.01 | -0.33 | 0.02 | -0.4^1^ | **5.4×10^-21^** | - | - | - | - | - | - | - |
|  | rs6496608 | A | *ANPEP* | -0.02 | 0.83 | 0.01 | 0.96 | - | **-** | - | - | - | - | - | - | - |
|  | **rs11073891** | C | *ANPEP* | -0.07 | 0.19 | -0.03 | 0.56 | -108.6 | **4.2×10^-302^** | - | - | - | - | - | - | 34 |
|  |  |  | *ARPIN* | -0.07 | 0.02 | 0.09 | 0.16 | 18.09 | **4.5×10^-65^** | - | - | - | - | - | - | - |
|  |  |  | *AP3S2* | -0.22 | **0.00002** | -0.22 | **9.2×10^-6^** | 7.85 | **5.5×10^-7^** | - | - | - | - | 0.24 | **3.7×10^-6^** | - |
|  |  |  | *IDH2* | -0.03 | 0.36 | -0.01 | 0.92 | 5.73 | **0.00006** | - | - | - | - | - | - | - |
|  |  |  | *MESP1* | 0.02 | 0.78 | 0.01 | 0.88 | 4.85 | **0.003** | - | - | - | - | - | - | - |
| rs12148357 | | T | *ANPEP* | -0.02 | 0.65 | 0.08 | 0.13 | - | - | - | - | - | - | - | - | 3 |
|  |  |  | *AP3S2* | -0.34 | **1.2×10^-12^** | -0.39 | **1.3×10^-8^** | -48.68 | **4.2×10^-302^** | - | - | - | - | 0.30 | **1.5×10^-8^** | - |
|  |  |  | *ARPIN* | 0.03 | 0.32 | 0.02 | 0.75 | -9.12 | **9.5×10^-12^** | - | - | - | - | - | - | - |
|  |  |  | *U7* | NA | NA | NA | NA | -10.51 | **9.8×10^-18^** | - | - | - | - | - | - | - |
|  |  |  | *PEX11A* | 0.01 | 0.90 | -0.02 | 0.74 | -4.51 | **0.02** | - | - | - | - | - | - | - |
| Haplotype block 4 | rs9920421 | G | *ANPEP* | -0.12 (A) | 0.03 | 0.02 (A) | 0.74 | 49.26 | **4.2×10^-302^** | - | - | - | - | - | - | 12 |
|  |  |  | *AP3S2* | -0.61 (A) | **4.7×10^-45^** | -0.67 (A) | **3.8×10^-24^** | 51.64 | **4.2×10^-302^** | -0.71 | **2.0×10^-15^** | -0.58 | **2.3×10^-7^** | 0.76 | **4.7×10^-47^** | - |
|  |  |  | *ARPIN* | -0.02 (A) | 0.52 | 0.07 (A) | 0.31 | -27.42 | **1.7×10^-157^** | 0.43 | **4.2×10^-8^** | - | - | - | - | - |
|  |  |  | *MESP1* | 0.09 | 0.08 | 0.07 | 0.38 | -4.66 | **0.008** | - | - | - | - | - | - | - |
|  |  |  | *U7* | NA | NA | NA | NA | 4.54 | **0.01** | - | - | - | - | - | - | - |
|  | **rs4932143** | G | *ANPEP* | -0.12 | 0.03 | 0.01 | 0.88 | 48.6 | **4.2×10^-302^** | - | - | - | - | - | - | 12 |
|  |  |  | *AP3S2* | -0.65 (C) | **1.5×10^-48^** | -0.69 (C) | **3.6×10^-25^** | 52.8 | **4.2×10^-302^** | -0.75 | **1.6×10^-16^** | -0.67 | **6.2×10^-9^** | 0.78 | **6.9×10^-48^** | - |
|  |  |  | *ARPIN* | -0.02 (C) | 0.61 | 0.05 (C) | 0.51 | -27.7 | **8.0×10^-161^** | 0.47 | **9.2×10^-9^** | - | - | - | - | - |
|  |  |  | *MESP1* | 0.10 | 0.09 | 0.04 | 0.64 | -4.9 | **0.003** | - | - | - | - | - | - | - |
|  |  |  | *U7* | NA | NA | NA | NA | 4.7 | **0.007** | - | - | - | - | - | - | - |
|  | rs7111 | T | *ANPEP* | -0.13 | 0.02 | -0.01 | 0.94 | 48.76 | **4.2×10^-302^** | - | - | - | - | - | - | 13 |
|  |  |  | *AP3S2* | -0.65 (C) | **2.4×10^-51^** | -0.74(C) | **2.3×10^-29^** | 53.17 | **4.2×10^-302^** | -0.75 | **8.1×10^-17^** | -0.66 | **1.6×10^-8^** | 0.78 | **1.5×10^-49^** | - |
|  |  |  | *ARPIN* | -0.01 (C) | - 0.83 | 0.06 (C) | 0.39 | -27.31 | **3.7×10^-156^** | 0.47 | **2.9×10^-9^** | - | - | - | - | - |
|  |  |  | *MESP1* | 0.07 | 0.22 | 0.04 | 0.62 | -5.12 | **9.6×10^-4^** | - | - | - | - | - | - | - |
|  |  |  | *U7* | NA | NA | NA | NA | 4.99 | **0.002** | - | - | - | - | - | - | - |
| rs72756574 | | G | *ANPEP* | -0.07 | 0.50 | -0.16 | 0.14 | -32.41 | **2.4×10^-222^** | 0.49 | **1.2×10^-7^** | - | - | 0.37 | **6.3×10^-6^** | - |
|  |  |  | *ARPIN* | -0.03 | 0.63 | 0.40 | 0.002 | 11.78 | **6.6×10^-24^** | - | - | - | - | - | - | - |
|  |  |  | *AP3S2* | -0.10 | 0.28 | -0.05 | 0.73 | 8.14 | **5.1×10^-8^** | - | - | - | - | - | - | - |
|  |  |  | *MESP1* | -0.00 | 0.97 | 0.07 | 0.65 | 4.94 | **0.002** | - | - | - | - | - | - | - |
| ^1^Data from Genotype-Tissue Expression (GTEx) project (https://www.gtexportal.org/home/);^2^Data from eQTLGen Consortium (https://www.eqtlgen.org/phase1.html/);^2^Data the number of mQTL from QTLbase2 (http://www.mulinlab.org/qtlbase). SNPs associated with T2D risk are bolded. | | | | | | | | | | | | | | | | |
