## Supplemental Table J for "The link between the *ANPEP* gene and type 2 diabetes mellitus may be mediated by the disruption of glutathione metabolism and redox homeostasis"

**Supplementary Table J**

Chromatin states and histone marks in pancreatic islets, pancreas and liver for the *ANPEP* polymorphisms associated with type 2 diabetes mellitus*

| Tissues/organ | Chromatin states | | Histone marks | | | | DNase |
| --- | --- | --- | --- | --- | --- | --- | --- |
|  | 15-state core chromatin model across tissues | 25-state core chromatin model across tissues | H3K4me1 | H3K4me3 | H3K27ac | H3K9ac |  |
| **rs25653 (no variants with r^2^ >= 0.8)** | | | | | | | |
| Pancreatic islets | 1_TssA |  |  | H3K4me3_Pro |  |  |  |
| Pancreas | 1_TssA | 9_TxReg | H3K4me1_Enh | H3K4me3_Pro | H3K27ac_Enh |  | DNase |
| Liver | 2_TssAFlnk | 9_TxReg | H3K4me1_Enh | H3K4me3_Pro | H3K27ac_Enh | H3K9ac_Pro |  |
| **rs12898828 (no variants with r^2^ >= 0.8)** | | | | | | | |
| Pancreatic islets |  |  |  |  |  |  |  |
| Pancreas |  |  | H3K4me1_Enh |  |  |  |  |
| Liver |  |  |  |  |  |  |  |
| **rs11073891 (4 variants with r^2^ >= 0.8)** | | | | | | | |
| Pancreatic islets |  |  |  |  |  |  |  |
| Pancreas |  |  |  |  |  |  |  |
| Liver |  |  |  |  | H3K27ac_Enh |  |  |
| **rs12148357 (out of haplotypes)** | | | | | | | |
| Pancreatic islets |  |  |  |  |  |  |  |
| Pancreas |  |  |  |  |  |  |  |
| Liver |  | 17_EnhW2 |  |  | H3K27ac_Enh |  |  |
| **rs9920421 (28 variants with r^2^ >= 0.8)** | | | | | | | |
| Pancreatic islets |  |  |  |  |  |  |  |
| Pancreas |  |  | H3K4me1_Enh |  |  |  |  |
| Liver |  |  |  |  | H3K27ac_Enh |  |  |
| **rs4932143 (45 variants with r^2^ >= 0.8)** | | | | | | | |
| Pancreatic islets |  | 17_EnhW2 |  |  |  |  |  |
| Pancreas | 7_Enh | 14_EnhA2 | H3K4me1_Enh |  | H3K27ac_Enh |  | DNase |
| Liver |  |  |  |  |  |  |  |
| **rs7111 (45 variants with r^2^ >= 0.8)** | | | | | | | |
| Pancreatic islets |  |  |  |  |  |  |  |
| Pancreas |  |  |  |  |  |  | DNase |
| Liver |  |  | H3K4me1_Enh |  | H3K27ac_Enh | H3K9ac_Pro |  |
| *Data of the Roadmap Epigenomics project from the HaploReg v4.2 online tool (https://pubs.broadinstitute.org/mammals/haploreg/haploreg.php)Chromatin States are as follows: 1_TssA, Active TSS (active transcription start sites); 2_TssAFlnk, Flanking active TSS; 9_TxReg, Transcribed & regulatory (Prom/Enh); 7_Enh, Enhancers; 17_EnhW2, Weak Enhancer 2; 14_EnhA2, Active Enhancer 2. Histone marks are as follows: H3K4me1_Enh and H3K27ac_Enh; enhancer-specific histone modifications; H3K4me3_Pro and H3K9ac_Pro, promoter-specific histone modifications; DNase, Primary DNase. | | | | | | | |
