## Supplemental Table E for "The link between the *ANPEP* gene and type 2 diabetes mellitus may be mediated by the disruption of glutathione metabolism and redox homeostasis"

**Supplementary Тable E**

*ANPEP* diplotypes significantly associated with the risk of type 2 diabetes

| № | Diplotypes | T2D | | Controls | | ^3^OR | CI- | CI+ | P | FDR |
| --- | --- | --- | --- | --- | --- | --- | --- | --- | --- | --- |
|  |  | n^1^ | %^2^ | n^1^ | %^2^ |  |  |  |  |  |
| Entire groups | | | | | | | | | | |
| 1 | rs9920421-A/G× rs4932143-С/C | 5 | 0,3 | 134 | 9,6 | 0,03 | 0,01 | 0,08 | 4,96E-32 | 5,65E-29 |
| 2 | rs12898828-A/A × rs4932143-C/C | 611 | 41,1 | 808 | 59,2 | 0,48 | 0,41 | 0,56 | 5,23E-22 | 3,97E-19 |
| 3 | rs17240268-G/G × rs4932143-C/C | 688 | 44,1 | 859 | 61,2 | 0,50 | 0,43 | 0,58 | 1,48E-20 | 8,42E-18 |
| 4 | rs41276922-G/G × rs4932143-C/C | 686 | 44,0 | 853 | 60,6 | 0,51 | 0,44 | 0,59 | 2,08E-19 | 9,47E-17 |
| 5 | rs8041622-G/G × rs4932143-C/C | 620 | 39,7 | 781 | 55,7 | 0,52 | 0,45 | 0,61 | 3,23E-18 | 1,23E-15 |
| 6 | rs6496608-T/T × rs4932143-C/C | 521 | 33,4 | 685 | 49,1 | 0,52 | 0,45 | 0,60 | 4,23E-18 | 1,38E-15 |
| 7 | rs4932143-C/C× rs72756574-T/T | 680 | 43,8 | 832 | 59,4 | 0,53 | 0,46 | 0,62 | 2,13E-17 | 5,59E-15 |
| 8 | rs72754570-G/G × rs4932143-C/C | 689 | 44,2 | 802 | 60,0 | 0,53 | 0,46 | 0,61 | 2,21E-17 | 5,59E-15 |
| 9 | rs4932143-G/G × rs7111-C/T | 611 | 39,9 | 345 | 25,5 | 1,94 | 1,65 | 2,27 | 2,89E-16 | 6,58E-14 |
| 10 | rs13380049-G/G × rs4932143-C/C | 702 | 45,2 | 841 | 60,2 | 0,54 | 0,47 | 0,63 | 3,35E-16 | 6,93E-14 |
| 11 | rs9920421-A/G× rs4932143-C/G | 618 | 39,9 | 361 | 25,9 | 1,90 | 1,62 | 2,22 | 9,31E-16 | 1,71E-13 |
| 12 | rs16974181-C/C × rs4932143-C/C | 564 | 37,9 | 715 | 52,9 | 0,54 | 0,47 | 0,63 | 9,79E-16 | 1,71E-13 |
| 13 | rs12148357-C/T× rs4932143-C/C | 331 | 21,7 | 469 | 34,1 | 0,53 | 0,45 | 0,63 | 7,22E-14 | 1,17E-11 |
| 14 | rs13380049-G/G × rs4932143-C/G | 612 | 39,4 | 382 | 27,3 | 1,73 | 1,48 | 2,02 | 4,90E-12 | 7,44E-10 |
| 15 | rs4932143-C/G× rs72756574-T/T | 604 | 38,9 | 378 | 27,0 | 1,72 | 1,47 | 2,01 | 7,41E-12 | 1,05E-09 |
| 16 | rs17240268-G/G × rs4932143-C/G | 600 | 38,5 | 376 | 26,8 | 1,71 | 1,46 | 2,00 | 1,41E-11 | 1,89E-09 |
| 17 | rs8041622-G/G × rs4932143-C/G | 587 | 37,6 | 367 | 26,2 | 1,70 | 1,45 | 1,99 | 2,99E-11 | 3,78E-09 |
| 18 | rs16974181-C/C × rs4932143-C/G | 514 | 34,5 | 314 | 23,2 | 1,74 | 1,48 | 2,06 | 3,68E-11 | 4,41E-09 |
| 19 | rs12442778-A/G × rs4932143-C/C | 395 | 25,6 | 494 | 36,6 | 0,59 | 0,51 | 0,70 | 1,28E-10 | 1,46E-08 |
| 20 | rs8034807-C/C × rs4932143-C/C | 393 | 25,3 | 499 | 36,1 | 0,60 | 0,51 | 0,70 | 2,06E-10 | 2,23E-08 |
| 21 | rs12898828-A/A × rs4932143-C/G | 562 | 37,8 | 366 | 26,8 | 1,66 | 1,42 | 1,95 | 3,72E-10 | 3,85E-08 |
| 22 | rs41276922-G/G × rs4932143-C/G | 550 | 35,3 | 349 | 24,8 | 1,66 | 1,41 | 1,94 | 4,91E-10 | 4,86E-08 |
| 23 | rs6496608-T/T × rs4932143-C/G | 529 | 33,9 | 337 | 24,2 | 1,61 | 1,37 | 1,89 | 6,08E-09 | 5,77E-07 |
| 24 | rs9920421-G/G× rs4932143-G/G | 118 | 7,6 | 39 | 2,8 | 2,86 | 1,98 | 4,14 | 6,42E-09 | 5,85E-07 |
| 25 | rs12148357-C/T× rs4932143-C/G | 389 | 25,5 | 229 | 16,7 | 1,71 | 1,42 | 2,05 | 6,83E-09 | 5,98E-07 |
| 26 | rs41276922-G/G × rs4932143-G/G | 107 | 6,9 | 34 | 2,4 | 2,98 | 2,01 | 4,41 | 1,26E-08 | 1,05E-06 |
| 27 | rs6496604-A/G × rs4932143-C/C | 349 | 24,4 | 440 | 34,3 | 0,62 | 0,52 | 0,73 | 1,29E-08 | 1,05E-06 |
| 28 | rs4932143-G/G× rs72756574-T/T | 124 | 8,0 | 44 | 3,1 | 2,67 | 1,88 | 3,80 | 1,41E-08 | 1,11E-06 |
| 29 | rs8041622-G/G× rs4932143-G/G | 122 | 7,8 | 43 | 3,1 | 2,68 | 1,88 | 3,82 | 1,83E-08 | 1,35E-06 |
| 30 | rs17240268-G/G × rs4932143-G/G | 115 | 7,4 | 39 | 2,8 | 2,79 | 1,92 | 4,04 | 1,84E-08 | 1,35E-06 |
| 31 | rs10152918-G/A × rs4932143-C/C | 381 | 24,8 | 478 | 34,2 | 0,63 | 0,54 | 0,74 | 1,93E-08 | 1,37E-06 |
| 32 | rs6496608-T/T× rs4932143-G/G | 122 | 7,8 | 43 | 3,1 | 2,67 | 1,87 | 3,81 | 2,14E-08 | 1,48E-06 |
| 33 | rs12148357-C/C× rs4932143-G/G | 105 | 6,9 | 34 | 2,5 | 2,91 | 1,96 | 4,32 | 2,91E-08 | 1,95E-06 |
| 34 | rs4932143-G/G × rs7111-T/T | 117 | 7,6 | 40 | 3,0 | 2,71 | 1,88 | 3,91 | 3,27E-08 | 2,13E-06 |
| 35 | rs72754570-G/G × rs4932143-C/G | 553 | 35,5 | 347 | 26,0 | 1,57 | 1,34 | 1,84 | 3,46E-08 | 2,19E-06 |
| 36 | rs11073889-T/C × rs4932143-C/C | 397 | 25,8 | 485 | 35,2 | 0,64 | 0,55 | 0,75 | 3,83E-08 | 2,35E-06 |
| 37 | rs13380049-G/G × rs4932143-G/G | 118 | 7,6 | 42 | 3,0 | 2,65 | 1,85 | 3,80 | 3,93E-08 | 2,35E-06 |
| 38 | rs12442778-A/G × rs4932143-C/G | 349 | 22,6 | 197 | 14,6 | 1,71 | 1,41 | 2,07 | 4,31E-08 | 2,52E-06 |
| 39 | rs12898828-A/A × rs4932143-G/G | 120 | 8,1 | 45 | 3,3 | 2,58 | 1,81 | 3,66 | 4,80E-08 | 2,73E-06 |
| 40 | rs6496604-A/G × rs4932143-C/G | 292 | 20,4 | 162 | 12,6 | 1,77 | 1,44 | 2,18 | 6,23E-08 | 3,46E-06 |
| 41 | rs6496603-A/G × rs4932143-C/G | 302 | 19,4 | 168 | 12,1 | 1,75 | 1,42 | 2,14 | 6,93E-08 | 3,76E-06 |
| 42 | rs72754570-G/G× rs4932143-G/G | 105 | 6,7 | 33 | 2,5 | 2,85 | 1,92 | 4,25 | 7,74E-08 | 4,10E-06 |
| 43 | rs11073889-T/C × rs4932143-C/G | 327 | 21,2 | 187 | 13,6 | 1,72 | 1,41 | 2,09 | 7,93E-08 | 4,10E-06 |
| 44 | rs12442778-A/A × rs4932143-G/G | 107 | 6,9 | 36 | 2,7 | 2,71 | 1,85 | 3,99 | 1,36E-07 | 6,88E-06 |
| 45 | rs753362-C/C × rs4932143-C/G | 321 | 20,9 | 183 | 13,5 | 1,70 | 1,39 | 2,07 | 1,47E-07 | 7,28E-06 |
| 46 | rs11073891-C/C × rs4932143-C/C | 224 | 15,5 | 244 | 23,9 | 0,58 | 0,48 | 0,72 | 1,63E-07 | 7,90E-06 |
| 47 | rs8041622-G/G × rs11073891-A/A | 369 | 25,5 | 184 | 16,8 | 1,69 | 1,38 | 2,05 | 1,86E-07 | 8,82E-06 |
| 48 | rs12442778-A/G × rs11073891-C/C | 9 | 0,6 | 37 | 3,5 | 0,18 | 0,09 | 0,37 | 2,63E-07 | 1,20E-05 |
| 49 | rs4932143-G/G × rs7111-T/T | 7 | 0,5 | 40 | 3,0 | 0,16 | 0,07 | 0,35 | 2,64E-07 | 1,20E-05 |
| 50 | rs11073891-A/A× rs72756574-T/T | 450 | 31,2 | 241 | 22,0 | 1,61 | 1,34 | 1,92 | 2,70E-07 | 1,21E-05 |
| 51 | rs16943590-C/C× rs4932143-C/C | 321 | 21,3 | 387 | 29,7 | 0,64 | 0,54 | 0,76 | 3,18E-07 | 1,39E-05 |
| 52 | rs16974181-C/C × rs4932143-G/G | 104 | 7,0 | 38 | 2,8 | 2,60 | 1,78 | 3,79 | 3,42E-07 | 1,47E-05 |
| 53 | rs16974181-C/C× rs11073891-A/A | 332 | 24,0 | 167 | 15,7 | 1,69 | 1,38 | 2,08 | 4,88E-07 | 2,02E-05 |
| 54 | rs6496603-A/A× rs4932143-C/C | 272 | 17,5 | 347 | 25,0 | 0,63 | 0,53 | 0,76 | 5,45E-07 | 2,22E-05 |
| 55 | rs17240268-G/G × rs11073891-C/C | 226 | 15,6 | 257 | 23,4 | 0,61 | 0,50 | 0,74 | 7,20E-07 | 2,88E-05 |
| 56 | rs8041622-G/G × rs11073891-C/C | 234 | 16,1 | 262 | 24,0 | 0,61 | 0,50 | 0,74 | 8,02E-07 | 3,15E-05 |
| 57 | rs72754570-G/G × rs11073891-C/C | 225 | 15,5 | 243 | 23,3 | 0,60 | 0,49 | 0,74 | 9,42E-07 | 3,64E-05 |
| 58 | rs41276922-G/G × rs11073891-C/C | 224 | 15,5 | 254 | 23,1 | 0,61 | 0,50 | 0,75 | 1,10E-06 | 4,17E-05 |
| 59 | rs12898828-A/A× rs11073891-A/A | 356 | 25,4 | 187 | 17,3 | 1,63 | 1,34 | 1,99 | 1,16E-06 | 4,33E-05 |
| 60 | rs25653-C/C × rs4932143-C/C | 171 | 11,1 | 234 | 17,2 | 0,60 | 0,48 | 0,74 | 1,63E-06 | 5,99E-05 |
| 61 | rs9920421-G/G× rs4932143-C/G | 8 | 0,5 | 39 | 2,8 | 0,19 | 0,09 | 0,40 | 1,71E-06 | 6,18E-05 |
| 62 | rs12898828-A/A × rs11073891-C/C | 234 | 16,7 | 264 | 24,4 | 0,62 | 0,51 | 0,76 | 2,10E-06 | 7,47E-05 |
| 63 | rs25653-T/C × rs4932143-C/G | 336 | 21,7 | 202 | 14,9 | 1,59 | 1,31 | 1,92 | 2,15E-06 | 7,53E-05 |
| 64 | rs16974181-C/C × rs11073891-C/C | 206 | 14,9 | 237 | 22,3 | 0,61 | 0,50 | 0,75 | 2,31E-06 | 7,97E-05 |
| 65 | rs13380049-G/G× rs11073891-A/A | 412 | 28,5 | 223 | 20,4 | 1,55 | 1,29 | 1,87 | 3,23E-06 | 0,0001 |
| 66 | rs10152918-G/A × rs4932143-G/G | 61 | 4,0 | 17 | 1,2 | 3,35 | 1,95 | 5,77 | 3,75E-06 | 0,0001 |
| 67 | rs12442778-A/A× rs11073891-A/A | 449 | 31,3 | 240 | 22,9 | 1,53 | 1,28 | 1,84 | 4,02E-06 | 0,0001 |
| 68 | rs753362-C/C × rs4932143-C/C | 376 | 24,5 | 437 | 32,1 | 0,68 | 0,58 | 0,80 | 4,31E-06 | 0,0001 |
| 69 | rs8034807-C/A × rs4932143-C/G | 250 | 16,1 | 143 | 10,3 | 1,66 | 1,33 | 2,07 | 5,10E-06 | 0,0002 |
| 70 | rs25653-T/C × rs4932143-G/G | 62 | 4,0 | 17 | 1,3 | 3,29 | 1,92 | 5,66 | 5,19E-06 | 0,0002 |
| 71 | rs17240268-G/G× rs11073891-A/A | 409 | 28,2 | 224 | 20,4 | 1,54 | 1,28 | 1,85 | 5,34E-06 | 0,0002 |
| 72 | rs8034807-C/C × rs4932143-G/G | 72 | 4,6 | 23 | 1,7 | 2,87 | 1,79 | 4,62 | 5,66E-06 | 0,0002 |
| 73 | rs753362-C/G× rs4932143-C/C | 334 | 21,7 | 395 | 29,0 | 0,68 | 0,57 | 0,80 | 5,71E-06 | 0,0002 |
| 74 | rs6496608-T/T × rs11073891-C/C | 251 | 17,3 | 267 | 24,5 | 0,65 | 0,53 | 0,78 | 8,87E-06 | 0,0003 |
| 75 | rs41276922-G/G × rs11073891-A/A | 386 | 26,7 | 211 | 19,2 | 1,53 | 1,27 | 1,85 | 9,76E-06 | 0,0003 |
| 76 | rs11073891-A/A× rs9920421-G/G | 89 | 6,2 | 27 | 2,5 | 2,60 | 1,68 | 4,03 | 9,82E-06 | 0,0003 |
| 77 | rs16943590-C/C× rs4932143-C/G | 316 | 21,0 | 190 | 14,6 | 1,55 | 1,28 | 1,89 | 1,11E-05 | 0,0003 |
| 78 | rs6496603-A/G× rs4932143-C/C | 383 | 24,6 | 442 | 31,9 | 0,70 | 0,59 | 0,82 | 1,22E-05 | 0,0004 |
| 79 | rs8034807-C/C × rs4932143-C/G | 350 | 22,5 | 224 | 16,2 | 1,50 | 1,25 | 1,81 | 1,65E-05 | 0,0005 |
| 80 | rs72754570-G/G × rs11073891-A/A | 385 | 26,6 | 200 | 19,2 | 1,52 | 1,26 | 1,85 | 1,76E-05 | 0,0005 |
| 81 | rs11073891-A/A× rs12148357-C/C | 155 | 10,9 | 65 | 6,0 | 1,91 | 1,41 | 2,58 | 1,96E-05 | 0,0005 |
| 82 | rs12442778-A/A × rs4932143-C/C | 175 | 11,3 | 227 | 16,8 | 0,63 | 0,51 | 0,78 | 1,97E-05 | 0,0005 |
| 83 | rs25653-T/C× rs4932143-C/C | 410 | 26,5 | 458 | 33,7 | 0,71 | 0,60 | 0,83 | 2,22E-05 | 0,0006 |
| 84 | rs11073891-C/C× rs72756574-T/T | 191 | 13,2 | 212 | 19,4 | 0,64 | 0,51 | 0,79 | 2,90E-05 | 0,0008 |
| 85 | rs4932143-C/C× rs7111-C/C | 778 | 50,8 | 791 | 58,5 | 0,73 | 0,63 | 0,85 | 3,25E-05 | 0,0009 |
| 86 | rs11073891-C/C × rs9920421-A/A | 221 | 15,4 | 237 | 21,7 | 0,65 | 0,53 | 0,80 | 3,91E-05 | 0,001 |
| 87 | rs11073889-C/C × rs4932143-C/C | 134 | 8,7 | 186 | 13,5 | 0,61 | 0,48 | 0,77 | 4,40E-05 | 0,001 |
| 88 | rs11073891-C/C × rs7111-C/C | 221 | 15,5 | 229 | 21,9 | 0,65 | 0,53 | 0,80 | 4,71E-05 | 0,001 |
| 89 | rs25653-C/C × rs11073891-C/C | 57 | 4,0 | 82 | 7,7 | 0,49 | 0,35 | 0,70 | 5,03E-05 | 0,001 |
| 90 | rs9920421-A/A× rs4932143-C/C | 782 | 50,5 | 807 | 57,9 | 0,74 | 0,64 | 0,86 | 5,18E-05 | 0,001 |
| 91 | rs13380049-G/G × rs11073891-C/C | 234 | 16,2 | 246 | 22,6 | 0,66 | 0,54 | 0,81 | 5,43E-05 | 0,001 |
| 92 | rs6496608-T/T × rs11073891-A/A | 280 | 19,3 | 145 | 13,3 | 1,56 | 1,25 | 1,94 | 5,81E-05 | 0,001 |
| 93 | rs16943590-C/C× rs4932143-G/G | 61 | 4,1 | 20 | 1,5 | 2,71 | 1,62 | 4,51 | 7,17E-05 | 0,002 |
| 94 | rs11073891-C/C × rs12148357-C/T | 107 | 7,5 | 132 | 12,2 | 0,58 | 0,45 | 0,76 | 7,45E-05 | 0,002 |
| 95 | rs753362-C/C × rs11073891-A/A | 234 | 16,4 | 115 | 10,9 | 1,61 | 1,27 | 2,05 | 8,15E-05 | 0,002 |
| 96 | rs6496604-A/A× rs4932143-C/C | 154 | 10,8 | 203 | 15,8 | 0,64 | 0,51 | 0,80 | 9,53E-05 | 0,002 |
| 97 | rs11073891-A/C× rs4932143-C/C | 382 | 26,4 | 344 | 33,6 | 0,71 | 0,59 | 0,84 | 9,84E-05 | 0,002 |
| 98 | rs11073891-A/C × rs41276922-G/A | 340 | 23,5 | 175 | 17,1 | 1,49 | 1,21 | 1,82 | 0,0001 | 0,002 |
| 99 | rs10152918-G/A × rs4932143-C/G | 293 | 19,1 | 193 | 13,8 | 1,47 | 1,20 | 1,79 | 0,0001 | 0,003 |
| 100 | rs16943590-C/T× rs4932143-C/C | 318 | 21,1 | 355 | 27,2 | 0,71 | 0,60 | 0,85 | 0,0001 | 0,003 |
| 101 | rs16943590-C/T × rs4932143-C/G | 234 | 15,5 | 139 | 10,7 | 1,54 | 1,23 | 1,93 | 0,0002 | 0,003 |
| 102 | rs753362-C/G × rs4932143-G/G | 54 | 3,5 | 18 | 1,3 | 2,71 | 1,58 | 4,65 | 0,0002 | 0,004 |
| 103 | rs753362-C/C × rs4932143-G/G | 58 | 3,8 | 21 | 1,5 | 2,50 | 1,51 | 4,14 | 0,0002 | 0,005 |
| 104 | rs11073891-A/A× rs7111-T/T | 86 | 6,0 | 30 | 2,9 | 2,17 | 1,42 | 3,32 | 0,0002 | 0,005 |
| 105 | rs8034807-C/C× rs11073891-A/A | 219 | 15,2 | 111 | 10,3 | 1,56 | 1,22 | 1,99 | 0,0003 | 0,006 |
| 106 | rs6496603-A/G × rs4932143-G/G | 59 | 3,8 | 22 | 1,6 | 2,45 | 1,49 | 4,01 | 0,0003 | 0,006 |
| 107 | rs6496604-A/G × rs4932143-G/G | 55 | 3,8 | 20 | 1,6 | 2,52 | 1,50 | 4,23 | 0,0003 | 0,006 |
| 108 | rs6496603-A/A× rs4932143-G/G | 53 | 3,4 | 19 | 1,4 | 2,54 | 1,50 | 4,31 | 0,0004 | 0,008 |
| 109 | rs10152918-G/A× rs11073891-A/A | 209 | 14,7 | 108 | 9,9 | 1,56 | 1,21 | 1,99 | 0,0004 | 0,008 |
| 110 | rs25653-T/T× rs11073891-A/A | 161 | 11,2 | 75 | 7,1 | 1,66 | 1,25 | 2,21 | 0,0005 | 0,01 |
| 111 | rs11073889-T/C × rs11073891-A/A | 228 | 15,9 | 119 | 11,0 | 1,53 | 1,20 | 1,93 | 0,0005 | 0,01 |
| 112 | rs6496604-A/G× rs11073891-A/A | 217 | 15,9 | 113 | 11,1 | 1,52 | 1,20 | 1,94 | 0,0006 | 0,01 |
| 113 | rs11073889-C/C × rs4932143-G/G | 40 | 2,6 | 12 | 0,9 | 3,03 | 1,58 | 5,81 | 0,0007 | 0,01 |
| 114 | rs10152918-G/G× rs4932143-C/G | 242 | 15,7 | 160 | 11,5 | 1,44 | 1,17 | 1,79 | 0,0008 | 0,02 |
| 115 | rs753362-C/G × rs4932143-C/G | 268 | 17,4 | 176 | 12,9 | 1,42 | 1,16 | 1,74 | 0,0008 | 0,02 |
| 116 | rs12442778-A/A × rs9920421-G/G | 111 | 7,2 | 66 | 4,4 | 1,69 | 1,24 | 2,32 | 0,0009 | 0,02 |
| 117 | rs10152918-A/A× rs4932143-C/G | 99 | 6,4 | 52 | 3,7 | 1,78 | 1,26 | 2,51 | 0,0009 | 0,02 |
| 118 | rs8034807-C/C × rs11073891-C/C | 142 | 9,8 | 152 | 14,1 | 0,67 | 0,52 | 0,85 | 0,001 | 0,02 |
| 119 | rs25653-T/T× rs4932143-C/G | 201 | 13,0 | 124 | 9,1 | 1,49 | 1,17 | 1,88 | 0,001 | 0,02 |
| 120 | rs11073889-T/C × rs4932143-G/G | 63 | 4,1 | 27 | 2,0 | 2,13 | 1,35 | 3,37 | 0,001 | 0,02 |
| 121 | rs753362-C/C× rs12898828-A/T | 118 | 8,1 | 76 | 5,1 | 1,63 | 1,21 | 2,19 | 0,001 | 0,02 |
| 122 | rs25653-C/C× rs12442778-A/G | 134 | 8,7 | 181 | 12,3 | 0,68 | 0,54 | 0,86 | 0,001 | 0,02 |
| 123 | rs8034807-C/A × rs4932143-C/C | 336 | 21,6 | 368 | 26,6 | 0,76 | 0,64 | 0,90 | 0,002 | 0,03 |
| 124 | rs25653-C/C × rs6496608-T/T | 227 | 14,7 | 289 | 19,0 | 0,73 | 0,61 | 0,89 | 0,002 | 0,03 |
| 125 | rs16943590-C/C× rs11073891-A/A | 181 | 12,9 | 90 | 8,9 | 1,53 | 1,17 | 2,00 | 0,002 | 0,03 |
| 126 | rs25653-T/T× rs4932143-G/G | 47 | 3,0 | 18 | 1,3 | 2,33 | 1,35 | 4,04 | 0,002 | 0,03 |
| 127 | rs6496604-G/G× rs6496608-A/A | 1 | 0,1 | 14 | 1,0 | 0,10 | 0,02 | 0,55 | 0,002 | 0,03 |
| 128 | rs6496608-T/A× rs11073891-A/A | 156 | 10,8 | 78 | 7,2 | 1,57 | 1,18 | 2,08 | 0,002 | 0,03 |
| 129 | rs10152918-G/G× rs4932143-C/C | 290 | 18,9 | 328 | 23,5 | 0,76 | 0,63 | 0,90 | 0,002 | 0,04 |
| 130 | rs25653-C/C × rs12898828-A/A | 224 | 15,2 | 289 | 19,5 | 0,74 | 0,61 | 0,90 | 0,002 | 0,04 |
| 131 | rs72754570-G/G × rs25653-C/C | 288 | 18,6 | 335 | 23,1 | 0,76 | 0,64 | 0,91 | 0,002 | 0,04 |
| 132 | rs8034807-C/A × rs4932143-G/G | 44 | 2,8 | 17 | 1,2 | 2,34 | 1,33 | 4,12 | 0,002 | 0,04 |
| 133 | rs6496603-A/A× rs16943590-C/T | 1 | 0,1 | 13 | 0,9 | 0,11 | 0,02 | 0,58 | 0,003 | 0,04 |
| 134 | rs12442778-A/G× rs12898828-A/A | 633 | 43,0 | 716 | 48,5 | 0,80 | 0,69 | 0,92 | 0,003 | 0,04 |
| 135 | rs12442778-A/G × rs6496608-T/T | 589 | 38,1 | 653 | 43,4 | 0,80 | 0,69 | 0,93 | 0,003 | 0,05 |
| 136 | rs8034807-C/A× rs6496608-T/A | 189 | 12,1 | 138 | 8,9 | 1,41 | 1,12 | 1,78 | 0,003 | 0,05 |
| Males | | | | | | | | | | |
| 1 | rs9920421-A/G × rs4932143-C/C | 2 | 0,3 | 52 | 9,9 | 0,04 | 0,01 | 0,14 | 5,2E-13 | 1,2E-09 |
| 2 | rs12898828-A/A × rs4932143-C/C | 246 | 43,6 | 307 | 59,3 | 0,53 | 0,42 | 0,68 | 2,7E-07 | 2,0E-04 |
| 3 | rs8041622-G/G × rs4932143-C/C | 249 | 42,4 | 300 | 56,6 | 0,56 | 0,45 | 0,72 | 2,2E-06 | 1,0E-03 |
| 4 | rs17240268-G/G × rs4932143-C/C | 281 | 47,9 | 328 | 61,9 | 0,57 | 0,45 | 0,72 | 2,6E-06 | 1,0E-03 |
| 5 | rs4932143-C/C × rs72756574-T/T | 273 | 46,9 | 323 | 60,9 | 0,57 | 0,45 | 0,72 | 2,8E-06 | 1,0E-03 |
| 6 | rs41276922-G/G × rs4932143-C/C | 280 | 47,8 | 327 | 61,6 | 0,57 | 0,45 | 0,72 | 3,8E-06 | 1,1E-03 |
| 7 | rs12148357-C/T × rs4932143-C/C | 122 | 21,1 | 174 | 33,5 | 0,53 | 0,41 | 0,70 | 4,0E-06 | 1,1E-03 |
| 8 | rs9920421-A/G × rs4932143-C/G | 219 | 37,6 | 132 | 25,1 | 1,81 | 1,39 | 2,34 | 6,8E-06 | 1,7E-03 |
| 9 | rs13380049-G/G× rs4932143-C/C | 279 | 47,9 | 322 | 61,0 | 0,59 | 0,46 | 0,75 | 1,2E-05 | 2,6E-03 |
| 10 | rs16974181-C/C × rs4932143-C/C | 232 | 41,1 | 282 | 54,3 | 0,59 | 0,46 | 0,75 | 1,4E-05 | 2,8E-03 |
| 11 | rs6496608-T/T× rs4932143-C/C | 213 | 36,3 | 257 | 48,8 | 0,60 | 0,47 | 0,76 | 2,5E-05 | 4,5E-03 |
| 12 | rs17240268-G/G × rs4932143-C/G | 218 | 37,1 | 135 | 25,5 | 1,73 | 1,34 | 2,24 | 2,8E-05 | 4,5E-03 |
| 13 | rs72754570-G/G × rs4932143-C/C | 282 | 48,0 | 306 | 60,7 | 0,60 | 0,47 | 0,76 | 2,8E-05 | 4,5E-03 |
| 14 | rs4932143-C/G × rs7111-C/T | 213 | 37,2 | 131 | 25,5 | 1,73 | 1,33 | 2,25 | 3,5E-05 | 5,2E-03 |
| 15 | rs16943590-C/T× rs4932143-C/C | 115 | 20,3 | 154 | 31,0 | 0,57 | 0,43 | 0,75 | 5,7E-05 | 7,8E-03 |
| 16 | rs12898828-A/A × rs4932143-C/G | 210 | 37,2 | 134 | 25,9 | 1,70 | 1,31 | 2,21 | 6,1E-05 | 7,8E-03 |
| 17 | rs16974181-C/C× rs11073891-A/A | 128 | 24,9 | 58 | 14,2 | 2,00 | 1,42 | 2,81 | 6,3E-05 | 7,8E-03 |
| 18 | rs4932143-C/G × rs72756574-T/T | 217 | 37,3 | 139 | 26,2 | 1,67 | 1,29 | 2,16 | 7,9E-05 | 9,2E-03 |
| 19 | rs16974181-C/C × rs4932143-C/G | 185 | 32,8 | 115 | 22,2 | 1,71 | 1,31 | 2,25 | 9,2E-05 | 1,0E-02 |
| 20 | rs11073889-T/C× rs4932143-C/C | 145 | 25,4 | 191 | 36,3 | 0,60 | 0,46 | 0,78 | 9,6E-05 | 1,0E-02 |
| 21 | rs8041622-G/G × rs4932143-C/G | 211 | 36,0 | 134 | 25,3 | 1,66 | 1,28 | 2,15 | 1,2E-04 | 1,2E-02 |
| 22 | rs6496603-A/G× rs4932143-C/C | 138 | 23,8 | 181 | 34,2 | 0,60 | 0,46 | 0,78 | 1,3E-04 | 1,3E-02 |
| 23 | rs6496604-A/G × rs4932143-C/C | 133 | 24,5 | 168 | 35,2 | 0,60 | 0,46 | 0,79 | 2,0E-04 | 1,8E-02 |
| 24 | rs11073891-A/A × rs72756574-T/T | 169 | 31,8 | 87 | 21,0 | 1,76 | 1,31 | 2,37 | 2,0E-04 | 1,8E-02 |
| 25 | rs13380049-G/G × rs4932143-C/G | 219 | 37,6 | 144 | 27,3 | 1,60 | 1,24 | 2,07 | 2,6E-04 | 2,1E-02 |
| 26 | rs12148357-C/T × rs4932143-C/G | 136 | 23,6 | 77 | 14,8 | 1,77 | 1,30 | 2,41 | 2,6E-04 | 2,1E-02 |
| 27 | rs753362-C/C × rs4932143-C/G | 118 | 20,5 | 63 | 12,3 | 1,84 | 1,32 | 2,57 | 2,7E-04 | 2,1E-02 |
| 28 | rs72754570-G/G × rs11073891-A/A | 148 | 27,6 | 69 | 17,5 | 1,80 | 1,31 | 2,49 | 3,0E-04 | 2,2E-02 |
| 29 | rs41276922-G/G × rs11073891-A/A | 149 | 27,9 | 74 | 17,8 | 1,78 | 1,30 | 2,44 | 3,0E-04 | 2,2E-02 |
| 30 | rs12442778-A/G × rs4932143-C/C | 152 | 26,3 | 187 | 36,5 | 0,62 | 0,48 | 0,81 | 3,2E-04 | 2,3E-02 |
| 31 | rs41276922-C/G× rs4932143-C/G | 199 | 34,0 | 129 | 24,3 | 1,60 | 1,23 | 2,08 | 4,0E-04 | 2,8E-02 |
| 32 | rs17240268-G/G × rs11073891-A/A | 154 | 28,7 | 79 | 18,9 | 1,72 | 1,27 | 2,35 | 4,9E-04 | 3,1E-02 |
| 33 | rs16943590-T/T× rs10152918-G/G | 48 | 8,6 | 20 | 3,6 | 2,51 | 1,47 | 4,29 | 5,0E-04 | 3,1E-02 |
| 34 | rs10152918-G/G ×rs4932143-C/G | 88 | 15,3 | 45 | 8,5 | 1,95 | 1,33 | 2,85 | 5,0E-04 | 3,1E-02 |
| 35 | rs12442778-A/A× rs11073891-C/C | 168 | 31,9 | 87 | 21,6 | 1,70 | 1,26 | 2,29 | 5,0E-04 | 3,1E-02 |
| 36 | rs12442778-A/G × rs4932143-C/G | 121 | 21,0 | 67 | 13,1 | 1,77 | 1,28 | 2,45 | 5,6E-04 | 3,4E-02 |
| 37 | rs25653-C/C× rs11073891-C/C | 18 | 3,4 | 35 | 8,7 | 0,37 | 0,21 | 0,67 | 6,0E-04 | 3,5E-02 |
| 38 | rs11073891-C/C × rs12148357-C/G | 37 | 7,0 | 56 | 13,8 | 0,47 | 0,31 | 0,73 | 6,3E-04 | 3,6E-02 |
| Females | | | | | | | | | | |
| 1 | rs9920421-A/G × rs4932143-C/C | 3 | 0,3 | 82 | 9,5 | 0,03 | 0,01 | 0,10 | 3,7E-20 | 4,2E-17 |
| 2 | rs12898828-A/A × rs4932143-C/C | 365 | 39,6 | 501 | 59,2 | 0,45 | 0,37 | 0,55 | 2,0E-16 | 1,5E-13 |
| 3 | rs17240268-G/G × rs4932143-C/C | 407 | 41,8 | 531 | 60,8 | 0,46 | 0,39 | 0,56 | 4,6E-16 | 2,2E-13 |
| 4 | rs41276922-G/G × rs4932143-C/C | 406 | 41,8 | 526 | 60,0 | 0,48 | 0,40 | 0,58 | 5,3E-15 | 3,0E-12 |
| 5 | rs6496608-T/T × rs4932143-C/C | 308 | 31,7 | 428 | 49,3 | 0,48 | 0,39 | 0,58 | 1,2E-14 | 5,3E-12 |
| 6 | rs72754570-G/G × rs4932143-C/C | 407 | 41,9 | 496 | 59,6 | 0,49 | 0,40 | 0,59 | 6,7E-14 | 2,6E-11 |
| 7 | rs8041622-G/G × rs4932143-C/C | 371 | 38,1 | 481 | 55,2 | 0,50 | 0,42 | 0,60 | 2,1E-13 | 6,7E-11 |
| 8 | rs4932143-C/G × rs7111-C/T | 398 | 41,5 | 214 | 25,5 | 2,07 | 1,69 | 2,53 | 1,1E-12 | 3,0E-10 |
| 9 | rs4932143-C/C × rs72756574-T/T | 407 | 41,9 | 509 | 58,4 | 0,51 | 0,43 | 0,62 | 1,2E-12 | 3,1E-10 |
| 10 | rs13380049-G/G × rs4932143-C/C | 423 | 43,6 | 519 | 59,7 | 0,52 | 0,43 | 0,63 | 4,4E-12 | 1,0E-09 |
| 11 | rs16974181-C/C × rs4932143-C/C | 332 | 35,9 | 433 | 52,0 | 0,52 | 0,43 | 0,63 | 1,1E-11 | 2,3E-09 |
| 12 | rs9920421-A/G × rs4932143-C/G | 399 | 41,3 | 229 | 26,4 | 1,95 | 1,60 | 2,38 | 2,5E-11 | 4,7E-09 |
| 13 | rs8034807-C/C × rs4932143-C/C | 236 | 24,3 | 326 | 38,0 | 0,52 | 0,43 | 0,64 | 2,1E-10 | 3,8E-08 |
| 14 | rs13380049-G/G × rs4932143-C/G | 393 | 40,5 | 238 | 27,4 | 1,80 | 1,48 | 2,19 | 3,6E-09 | 5,7E-07 |
| 15 | rs12148357-C/T × rs4932143-C/C | 209 | 22,0 | 295 | 34,5 | 0,54 | 0,44 | 0,66 | 3,7E-09 | 5,7E-07 |
| 16 | rs4932143-C/G × rs72756574-T/T | 387 | 39,8 | 239 | 27,4 | 1,75 | 1,44 | 2,13 | 2,1E-08 | 3,1E-06 |
| 17 | rs9920421-G/G × rs4932143-G/G | 82 | 8,5 | 22 | 2,5 | 3,55 | 2,20 | 5,74 | 4,1E-08 | 5,5E-06 |
| 18 | rs8041622-G/G × rs4932143-G/G | 85 | 8,7 | 24 | 2,8 | 3,38 | 2,13 | 5,37 | 5,4E-08 | 6,2E-06 |
| 19 | rs13380049-G/G × rs4932143-G/G | 83 | 8,6 | 23 | 2,7 | 3,44 | 2,15 | 5,51 | 5,8E-08 | 6,2E-06 |
| 20 | rs8041622-G/G × rs4932143-C/G | 376 | 38,6 | 233 | 26,7 | 1,72 | 1,41 | 2,10 | 5,9E-08 | 6,2E-06 |
| 21 | rs17240268-G/G × rs4932143-G/G | 79 | 8,1 | 21 | 2,4 | 3,59 | 2,20 | 5,86 | 6,0E-08 | 6,2E-06 |
| 22 | rs6496608-T/T × rs4932143-G/G | 85 | 8,7 | 24 | 2,8 | 3,37 | 2,12 | 5,35 | 6,0E-08 | 6,2E-06 |
| 23 | rs16943590-C/C × rs4932143-C/C | 190 | 20,2 | 254 | 31,5 | 0,55 | 0,44 | 0,69 | 7,0E-08 | 6,7E-06 |
| 24 | rs4932143-G/G × rs72756574-T/T | 86 | 8,9 | 25 | 2,9 | 3,28 | 2,08 | 5,18 | 7,2E-08 | 6,7E-06 |
| 25 | rs12898828-A/A × rs4932143-G/G | 84 | 9,1 | 25 | 3,0 | 3,30 | 2,09 | 5,20 | 7,4E-08 | 6,7E-06 |
| 26 | rs12442778-A/G × rs4932143-C/C | 243 | 25,1 | 307 | 36,7 | 0,58 | 0,47 | 0,71 | 9,0E-08 | 7,7E-06 |
| 27 | rs41276922-G/G × rs4932143-G/G | 72 | 7,4 | 18 | 2,1 | 3,82 | 2,26 | 6,45 | 9,1E-08 | 7,7E-06 |
| 28 | rs16974181-C/C × rs4932143-C/G | 329 | 35,6 | 199 | 23,9 | 1,76 | 1,43 | 2,17 | 9,6E-08 | 7,8E-06 |
| 29 | rs17240268-G/G × rs4932143-C/G | 382 | 39,3 | 241 | 27,6 | 1,70 | 1,40 | 2,07 | 1,0E-07 | 7,9E-06 |
| 30 | rs4932143-G/G × rs7111-T/T | 81 | 8,5 | 22 | 2,6 | 3,42 | 2,12 | 5,53 | 1,2E-07 | 9,0E-06 |
| 31 | rs16974181-C/C × rs4932143-G/G | 74 | 8,0 | 20 | 2,4 | 3,53 | 2,14 | 5,85 | 1,9E-07 | 1,4E-05 |
| 32 | rs72754570-G/G × rs4932143-G/G | 71 | 7,3 | 17 | 2,0 | 3,78 | 2,21 | 6,47 | 2,3E-07 | 1,6E-05 |
| 33 | rs12148357-C/C × rs4932143-G/G | 73 | 7,7 | 20 | 2,3 | 3,48 | 2,10 | 5,76 | 2,8E-07 | 2,0E-05 |
| 34 | rs41276922-G/G × rs4932143-C/G | 351 | 36,1 | 220 | 25,1 | 1,69 | 1,38 | 2,06 | 3,0E-07 | 2,0E-05 |
| 35 | rs10152918-G/A × rs4932143-C/C | 235 | 24,4 | 304 | 35,1 | 0,60 | 0,49 | 0,73 | 5,9E-07 | 3,8E-05 |
| 36 | rs6496608-T/T × rs4932143-C/G | 338 | 34,7 | 210 | 24,2 | 1,67 | 1,36 | 2,04 | 7,8E-07 | 5,0E-05 |
| 37 | rs6496603-A/A× rs4932143-C/C | 163 | 16,7 | 224 | 26,2 | 0,57 | 0,45 | 0,71 | 8,2E-07 | 5,0E-05 |
| 38 | rs4932143-C/G × rs7111-T/T | 1 | 0,1 | 25 | 3,0 | 0,05 | 0,01 | 0,26 | 9,6E-07 | 5,7E-05 |
| 39 | rs12898828-A/A × rs4932143-C/G | 352 | 38,2 | 232 | 27,4 | 1,64 | 1,34 | 2,00 | 1,4E-06 | 8,4E-05 |
| 40 | rs12442778-A/A × rs4932143-G/G | 75 | 7,8 | 22 | 2,6 | 3,11 | 1,91 | 5,05 | 1,6E-06 | 8,8E-05 |
| 41 | rs10152918-G/A × rs4932143-C/G | 190 | 19,7 | 102 | 11,8 | 1,84 | 1,42 | 2,39 | 3,4E-06 | 1,9E-04 |
| 42 | rs11073891-C/C × rs4932143-C/C | 137 | 15,0 | 153 | 24,2 | 0,55 | 0,43 | 0,72 | 5,4E-06 | 2,9E-04 |
| 43 | rs6496604-A/G × rs4932143-C/G | 185 | 20,8 | 101 | 12,6 | 1,83 | 1,41 | 2,39 | 5,8E-06 | 3,1E-04 |
| 44 | rs12148357-C/T × rs4932143-C/G | 253 | 26,6 | 152 | 17,8 | 1,68 | 1,34 | 2,11 | 6,3E-06 | 3,3E-04 |
| 45 | rs72754570-G/G × rs4932143-C/G | 352 | 36,3 | 220 | 26,4 | 1,58 | 1,29 | 1,94 | 8,1E-06 | 4,1E-04 |
| 46 | rs11073889-T/C × rs4932143-C/G | 214 | 22,0 | 119 | 14,0 | 1,74 | 1,36 | 2,23 | 8,5E-06 | 4,2E-04 |
| 47 | rs10152918-G/A × rs4932143-G/G | 45 | 4,7 | 9 | 1,0 | 4,48 | 2,21 | 9,06 | 8,6E-06 | 4,2E-04 |
| 48 | rs9920421-G/G × rs4932143-C/G | 2 | 0,2 | 24 | 2,8 | 0,09 | 0,02 | 0,33 | 9,1E-06 | 4,3E-04 |
| 49 | rs6496603-A/G× rs4932143-C/G | 186 | 19,1 | 100 | 11,7 | 1,78 | 1,37 | 2,32 | 1,3E-05 | 6,0E-04 |
| 50 | rs12442778-A/G × rs11073891-C/C | 5 | 0,6 | 24 | 3,7 | 0,15 | 0,06 | 0,39 | 1,3E-05 | 6,0E-04 |
| 51 | rs12442778-A/A × rs4932143-C/C | 98 | 10,1 | 143 | 17,1 | 0,55 | 0,41 | 0,72 | 1,4E-05 | 6,2E-04 |
| 52 | rs6496604-A/G × rs4932143-C/C | 216 | 24,3 | 272 | 33,8 | 0,63 | 0,51 | 0,78 | 1,6E-05 | 7,1E-04 |
| 53 | rs8034807-C/C × rs4932143-G/G | 52 | 5,4 | 14 | 1,6 | 3,41 | 1,87 | 6,19 | 2,1E-05 | 8,9E-04 |
| 54 | rs12442778-A/G × rs4932143-C/G | 228 | 23,6 | 130 | 15,6 | 1,67 | 1,32 | 2,12 | 2,1E-05 | 9,0E-04 |
| 55 | rs25653-C/C × rs4932143-C/C | 103 | 10,6 | 147 | 17,4 | 0,56 | 0,43 | 0,74 | 2,7E-05 | 1,1E-03 |
| 56 | rs8041622-G/G × rs11073891-C/C | 139 | 15,2 | 159 | 23,5 | 0,58 | 0,45 | 0,75 | 2,9E-05 | 1,2E-03 |
| 57 | rs11073891-A/A × rs4932143-G/G | 63 | 6,9 | 14 | 2,2 | 3,27 | 1,82 | 5,89 | 3,2E-05 | 1,3E-03 |
| 58 | rs25653-T/C × rs4932143-C/C | 250 | 25,7 | 293 | 34,7 | 0,65 | 0,53 | 0,80 | 3,2E-05 | 1,3E-03 |
| 59 | rs4932143-C/C × rs7111-C/C | 468 | 48,8 | 491 | 58,6 | 0,67 | 0,56 | 0,81 | 3,3E-05 | 1,3E-03 |
| 60 | rs8041622-G/G × rs11073891-A/A | 238 | 26,0 | 117 | 17,3 | 1,69 | 1,31 | 2,16 | 3,4E-05 | 1,3E-03 |
| 61 | rs9920421-A/G × rs4932143-C/C | 470 | 48,6 | 503 | 58,1 | 0,68 | 0,57 | 0,82 | 4,9E-05 | 1,8E-03 |
| 62 | rs753362-C/C × rs4932143-C/C | 228 | 23,7 | 273 | 32,3 | 0,65 | 0,53 | 0,80 | 5,0E-05 | 1,8E-03 |
| 63 | rs17240268-G/G × rs11073891-C/C | 135 | 14,8 | 155 | 22,7 | 0,59 | 0,46 | 0,76 | 5,0E-05 | 1,8E-03 |
| 64 | rs16943590-C/C × rs4932143-G/G | 45 | 4,8 | 11 | 1,4 | 3,64 | 1,87 | 7,08 | 5,1E-05 | 1,8E-03 |
| 65 | rs41276922-G/G × rs11073891-C/C | 135 | 14,8 | 155 | 22,6 | 0,59 | 0,46 | 0,77 | 6,0E-05 | 2,1E-03 |
| 66 | rs11073889-T/C× rs4932143-C/C | 252 | 25,9 | 294 | 34,5 | 0,67 | 0,54 | 0,81 | 7,0E-05 | 2,4E-03 |
| 67 | rs12898828-A/A × rs11073891-A/A | 224 | 25,4 | 116 | 17,2 | 1,64 | 1,27 | 2,11 | 1,1E-04 | 3,7E-03 |
| 68 | rs6496603-A/A × rs4932143-G/G | 40 | 4,1 | 10 | 1,2 | 3,62 | 1,80 | 7,29 | 1,2E-04 | 4,0E-03 |
| 69 | rs753362-C/C × rs4932143-G/G | 42 | 4,4 | 11 | 1,3 | 3,47 | 1,77 | 6,78 | 1,2E-04 | 4,0E-03 |
| 70 | rs753362-C/C × rs4932143-C/G | 203 | 21,1 | 120 | 14,2 | 1,62 | 1,26 | 2,07 | 1,3E-04 | 4,2E-03 |
| 71 | rs753362-C/G× rs4932143-C/C | 208 | 21,6 | 249 | 29,4 | 0,66 | 0,53 | 0,82 | 1,4E-04 | 4,4E-03 |
| 72 | rs25653-T/C × rs4932143-G/G | 40 | 4,1 | 10 | 1,2 | 3,58 | 1,78 | 7,21 | 1,4E-04 | 4,4E-03 |
| 73 | rs12898828-A/A × rs11073891-C/C | 144 | 16,3 | 162 | 24,0 | 0,62 | 0,48 | 0,79 | 1,5E-04 | 4,7E-03 |
| 74 | rs16974181-C/C × rs11073891-C/C | 124 | 14,2 | 142 | 21,7 | 0,60 | 0,46 | 0,78 | 1,6E-04 | 4,9E-03 |
| 75 | rs11073891-A/A × rs12148357-C/C | 102 | 11,4 | 40 | 6,0 | 2,04 | 1,39 | 2,98 | 1,9E-04 | 5,8E-03 |
| 76 | rs13380049-G/G× rs11073891-A/A | 262 | 28,7 | 139 | 20,5 | 1,56 | 1,23 | 1,97 | 2,0E-04 | 5,9E-03 |
| 77 | rs72754570-G/G × rs11073891-C/C | 136 | 14,9 | 144 | 22,3 | 0,61 | 0,47 | 0,79 | 2,0E-04 | 5,9E-03 |
| 78 | rs16943590-C/C × rs4932143-C/G | 209 | 22,2 | 123 | 15,2 | 1,59 | 1,24 | 2,03 | 2,0E-04 | 6,0E-03 |
| 79 | rs25653-T/C × rs4932143-C/G | 212 | 21,8 | 127 | 15,0 | 1,58 | 1,24 | 2,01 | 2,1E-04 | 6,1E-03 |
| 80 | rs11073889-C/C × rs4932143-C/C | 74 | 7,6 | 109 | 12,8 | 0,56 | 0,41 | 0,77 | 2,5E-04 | 7,0E-03 |
| 81 | rs11073891-A/A × rs9920421-G/G | 61 | 6,7 | 18 | 2,7 | 2,63 | 1,54 | 4,50 | 2,5E-04 | 7,0E-03 |
| 82 | rs6496604-A/A × rs4932143-C/C | 89 | 10,0 | 128 | 15,9 | 0,59 | 0,44 | 0,79 | 2,9E-04 | 8,1E-03 |
| 83 | rs11073889-C/C × rs4932143-G/G | 31 | 3,2 | 6 | 0,7 | 4,36 | 1,87 | 10,20 | 3,0E-04 | 8,1E-03 |
| 84 | rs11073891-A/A × rs72756574-T/T | 281 | 30,8 | 154 | 22,7 | 1,52 | 1,21 | 1,91 | 3,0E-04 | 8,1E-03 |
| 85 | rs11073891-C/C × rs9920421-A/A | 135 | 14,9 | 148 | 21,9 | 0,62 | 0,48 | 0,81 | 3,1E-04 | 8,3E-03 |
| 86 | rs6496608-T/T × rs11073891-C/C | 154 | 16,9 | 163 | 24,1 | 0,64 | 0,50 | 0,82 | 3,5E-04 | 9,3E-03 |
| 87 | rs16943590-C/T× rs4932143-C/G | 146 | 15,5 | 80 | 9,9 | 1,67 | 1,25 | 2,23 | 4,9E-04 | 1,3E-02 |
| 88 | rs6496608-T/T × rs11073891-A/A | 179 | 19,6 | 88 | 13,0 | 1,63 | 1,23 | 2,15 | 5,2E-04 | 1,3E-02 |
| 89 | rs11073891-C/C × rs7111-C/C | 135 | 15,0 | 141 | 21,8 | 0,63 | 0,49 | 0,82 | 5,3E-04 | 1,4E-02 |
| 90 | rs8034807-C/C × rs4932143-C/G | 229 | 23,6 | 146 | 17,0 | 1,50 | 1,19 | 1,89 | 5,4E-04 | 1,4E-02 |
| 91 | rs8034807-C/A× rs4932143-C/G | 153 | 15,8 | 88 | 10,3 | 1,63 | 1,24 | 2,16 | 5,4E-04 | 1,4E-02 |
| 92 | rs12442778-A/A × rs9920421-G/G | 74 | 7,7 | 37 | 4,0 | 2,01 | 1,34 | 3,02 | 5,8E-04 | 1,4E-02 |
| 93 | rs13380049-G/G × rs11073891-C/C | 141 | 15,5 | 150 | 22,2 | 0,64 | 0,50 | 0,83 | 6,4E-04 | 1,6E-02 |
| 94 | rs753362-C/G × rs4932143-C/G | 174 | 18,1 | 104 | 12,3 | 1,58 | 1,21 | 2,05 | 6,5E-04 | 1,6E-02 |
| 95 | rs25653-T/T× rs4932143-G/G | 36 | 3,7 | 10 | 1,2 | 3,21 | 1,58 | 6,51 | 6,5E-04 | 1,6E-02 |
| 96 | rs10152918-G/A× rs12148357-C/C | 140 | 14,9 | 94 | 9,8 | 1,61 | 1,22 | 2,12 | 7,6E-04 | 1,8E-02 |
| 97 | rs16943590-T/T × rs4932143-C/C | 66 | 7,0 | 94 | 11,7 | 0,57 | 0,41 | 0,80 | 8,3E-04 | 1,9E-02 |
| 98 | rs753362-C/G × rs4932143-G/G | 34 | 3,5 | 9 | 1,1 | 3,28 | 1,59 | 6,76 | 1,0E-03 | 2,3E-02 |
| 99 | rs16974181-C/C × rs11073891-A/A | 204 | 23,4 | 109 | 16,6 | 1,53 | 1,19 | 1,99 | 1,1E-03 | 2,5E-02 |
| 100 | rs6496604-A/A × rs4932143-G/G | 31 | 3,5 | 8 | 1,0 | 3,44 | 1,60 | 7,39 | 1,1E-03 | 2,5E-02 |
| 101 | rs11073891-A/A × rs7111-T/T | 59 | 6,6 | 19 | 2,9 | 2,31 | 1,36 | 3,92 | 1,4E-03 | 3,2E-02 |
| 102 | rs12442778-A/G × rs6496608-T/T | 368 | 38,0 | 423 | 45,2 | 0,74 | 0,62 | 0,89 | 1,5E-03 | 3,3E-02 |
| 103 | rs12442778-A/A × rs11073891-A/A | 281 | 30,9 | 153 | 23,7 | 1,44 | 1,15 | 1,81 | 1,7E-03 | 3,7E-02 |
| 104 | rs12148357-C/C × rs9920421-G/G | 74 | 7,8 | 42 | 4,4 | 1,85 | 1,25 | 2,73 | 1,7E-03 | 3,7E-02 |
| 105 | rs8034807-C/C× rs12442778-A/G | 253 | 26,1 | 303 | 32,7 | 0,73 | 0,60 | 0,89 | 1,8E-03 | 3,9E-02 |
| 106 | rs11073889-C/C× rs10152918-G/G | 58 | 6,0 | 94 | 9,8 | 0,59 | 0,42 | 0,83 | 2,0E-03 | 4,3E-02 |
| 107 | rs11073891-A/C× rs4932143-C/G | 225 | 24,6 | 114 | 18,0 | 1,49 | 1,15 | 1,91 | 2,0E-03 | 4,3E-02 |
| 108 | rs6496604-A/G × rs4932143-G/G | 35 | 3,9 | 12 | 1,5 | 2,71 | 1,40 | 5,25 | 2,2E-03 | 4,6E-02 |
| 109 | rs17240268-G/G × rs11073891-A/A | 255 | 27,9 | 145 | 21,2 | 1,44 | 1,14 | 1,82 | 2,2E-03 | 4,6E-02 |
| ^1^Absolute number of individuals with particular diplotype. ^2^Percentage of individuals with diplotype. ^3^OR, odds ratio; 95% CI, confidence intervals.  P-values were adjusted for multiple testing by false discovery rate (FDR) using FDR online calculator (https://www.sdmproject.com/utilities/?show=FDR). | | | | | | | | | | |
